# Human-porcine transcriptomics reveals resuscitation-responsive pathways in trauma shock

**DOI:** 10.64898/2026.01.31.26345216

**Authors:** Jack S. Gisby, Robert Purcell, Isabella Withnell, Claudia P Cabrera, David S Watson, Sara Masarone, Gerard Hernández Mir, Emanuel Savage, Emma Bourne, Eva Wozniak, Charles A Mein, Jennifer Ross, Jason Pott, Joanna Shepherd, Daniel J Pennington, Sarah Watts, Emrys Kirkman, Karim Brohi, Michael R Barnes

## Abstract

Haemorrhage is the leading preventable cause of trauma death, primarily through ischaemic consequences that current treatments cannot adequately address. We combined human transcriptomic data (n=458) with a controlled porcine model of haemorrhagic shock to identify treatment-responsive molecular mechanisms. Using latent factorisation, we prioritised distinct molecular signatures of the human shock response, including stress signalling, neutrophil activation, and cytotoxic lymphocyte programmes. We assessed the behaviour of these pathways in the experimental porcine system, revealing that shock-initiated immune trajectories are not immutable: blood resuscitation normalised maladaptive transcriptomic changes whilst noradrenaline exacerbated them. While resuscitation modulated neutrophil, heat-shock, and barrier defence programmes, interferon and coagulation pathways were neither mortality-predictive nor treatment-responsive. A factor representing p38-MAPK/AP-1 stress signalling emerged as the dominant mortality-predictive pathway. An *in silico* small molecule screen identified p38 inhibitors as leading candidates for reversing shock-induced transcriptomic signatures. Our framework identifies modifiable pathways in trauma shock, prioritising p38-MAPK inhibition for therapeutic development and providing a systematic approach for trauma drug repurposing.

**One sentence summary**: Human-porcine cross-analysis reveals treatment-modifiable molecular signatures of trauma shock, identifying potential therapeutic strategies.

## Introduction

Trauma is a major and growing global health challenge, responsible for over 4 million deaths^1^ annually. Haemorrhage is the leading cause of preventable death in trauma^2,3^, and the key unmet needs in contemporary trauma care are treatments to mitigate the ischaemic consequences of shock. The depth and duration of trauma shock are key determinants of mortality across civilian and military arenas. Ischaemia drives immune dysregulation, organ dysfunction, multiple organ failure and death^4^. Recent advances in trauma care, particularly in resuscitation and haemostasis strategies, have stalled due to lack of options for shock management^5^. As such, there is a critical need for strategies that can support trauma patients during prolonged prehospital care, improving survival and reducing long-term complications^6^. Identifying effective therapeutic interventions to mitigate the effects of shock is an urgent priority^7^.

After injury, tissue damage and shock drive a complex cascade of immune and metabolic responses that directly influence clinical outcomes^8^. We and others have shown that there is a hyperacute immune response, occurring within minutes of injury, that cascades into a “genomic storm” involving a large portion of the whole blood transcriptome^9,10^. However, while trauma patients’ trajectories are set in the hyperacute phase after injury, it is unknown whether or how these transcriptomic changes can be modulated by treatment. Furthermore, it remains unknown whether dysregulated genes should be augmented or inhibited to optimise clinical outcomes.

Preclinical models of trauma and haemorrhagic shock provide controlled experimental systems for investigating the temporal dynamics of the injury response. Porcine models have emerged as particularly valuable for trauma research due to similarities in cardiovascular physiology, size, and coagulation systems^11–13^. Recently, such a model was employed to assess the effects of different fluid resuscitation strategies on the resolution of traumatic haemorrhagic shock, demonstrating that blood components are superior to saline when access to advanced medical care is delayed^14^. While these models offer clear potential for assessing therapeutic interventions, significant resources are required for their implementation and their translation to clinical practice requires careful consideration due to inherent species differences.

To address these challenges, we have adopted an integrated approach combining transcriptomic data from human trauma patients with a controlled porcine model of trauma and haemorrhagic shock. By comparing transcriptional networks associated with shock and mortality, we demonstrate strong conservation of core inflammatory and immune response networks between species. We show that blood product resuscitation effectively reverses shock-associated transcriptional changes, normalising maladaptive pathways, while noradrenaline exacerbates these responses, showing that immune trajectories initiated by trauma shock are not immutable and can be modified by clinical interventions. Using machine learning factorisation, we prioritise specific shock-induced transcriptional factors that can be exacerbated or resolved by different resuscitation strategies. We perform *in silico* transcriptomics-based screens to identify compounds that drive these signatures back towards normal, presenting therapeutic repositioning opportunities to improve outcomes in trauma care. These can in turn be tested within controlled experimental medicine frameworks^15^, enabling direct evaluation of candidate therapies under conditions that maintain both clinical realism and experimental control.

## Results

### Clinical characteristics of trauma patients and experimental model system

Of 2500 patients enrolled in the ACIT study^16^ between 2008 and 2018, we selected the 458 trauma patients without penetrating trauma or major traumatic brain injury, who had admission blood samples and complete data sets for analysis (**Fig. 1**). Patients had a spectrum of injury severity, with 30% graded as having a critical burden of injury (ISS >24; 138/458). Shock, defined as base deficit >6, was present in 48 patients (10%), and these had a higher median injury severity compared to non-shocked patients (29 vs 10) (**Table 1**). Mortality in shocked patients was 33% (16/48) compared to 3% (13/383) in the non-shocked group.

**Fig. 1.**
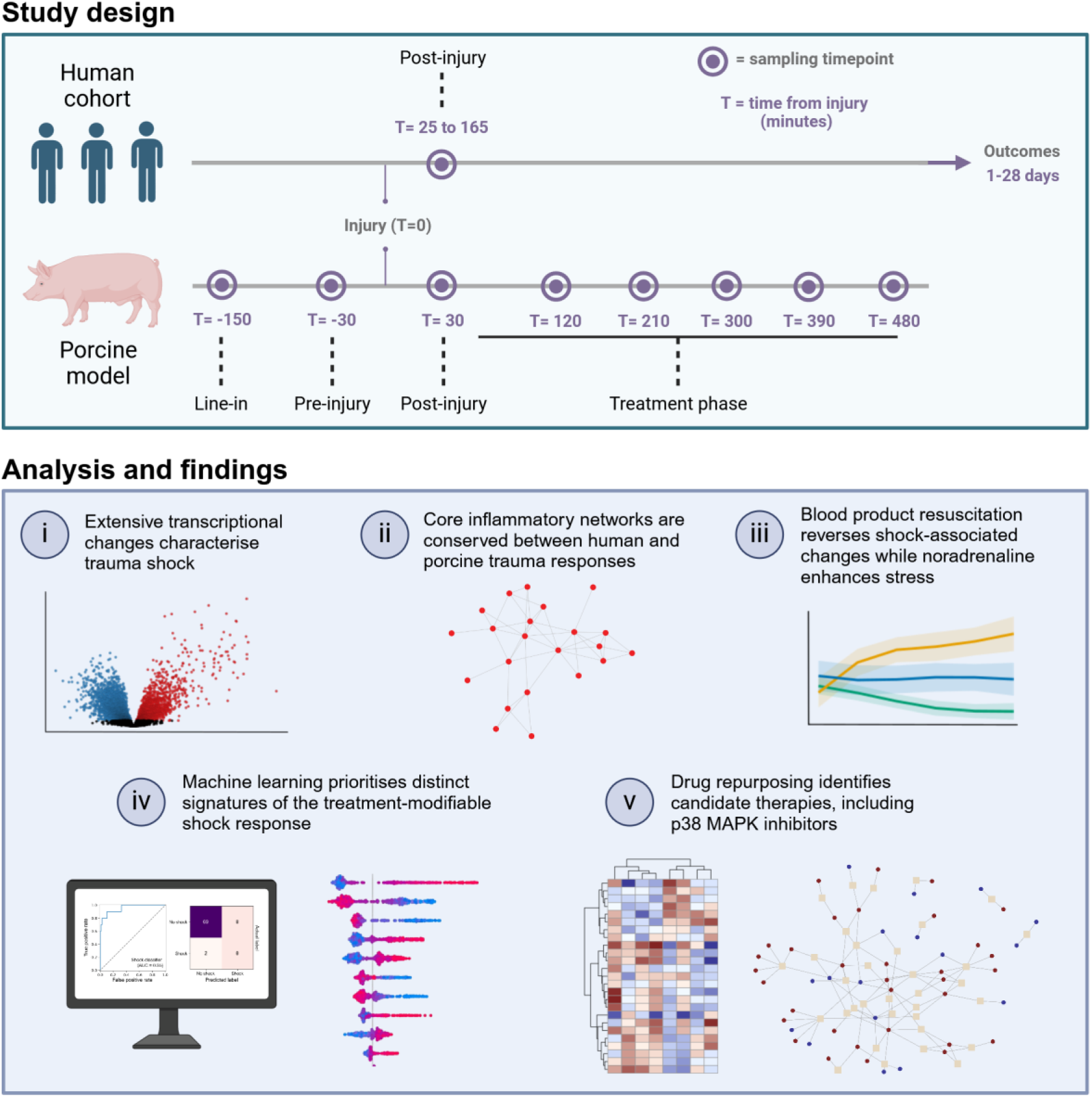
Study design and key analyses. Above, graphical summaries are shown for each of the two studies indicating the sampling procedure for each individual. Each human participant was sampled a single time immediately following injury. All patients were admitted within two hours post-injury and baseline samples were taken as soon as practicable. Each animal was sampled prior to surgery, prior to injury, prior to treatment, and repeatedly over time following treatment. Below, the major analyses and findings are highlighted.

**Table 1.** Baseline demographics of the cohort.

| Patient characteristic | Patient groups |  |  |
| --- | --- | --- | --- |
|  | All | No shock* | Shock* |
| **Count** | 458 | 383 | 48 |
| **Age** | 41.5 ± 17.3 | 41.0 ± 16.9 | 41.4 ± 17.6 |
| **Sex** |  |  |  |
| **Male** | 376 (82.1%) | 314 (82.0%) | 41 (85.4%) |
| **Female** | 82 (17.9%) | 69 (18.0%) | 7 (14.6%) |
| **Injury severity score** | 16.4 ± 13.4 | 15.2 ± 12.7 | 28.7 ± 13.8 |
| **Baseline measures** |  |  |  |
| **SBP (mmHg)** | 133.9 ± 28.6 | 135.5 ± 27.0 | 114.9 ± 34.8 |
| **WCC (x10<sup>9</sup>/L)** | 14.0 ± 6.8 | 13.7 ± 6.3 | 17.3 ± 9.3 |
| **Lactate (mmol/L)** | 2.6 ± 2.4 | 2.1 ± 1.3 | 6.8 ± 4.3 |
| **Base deficit (mmol/L)** | 1.6 ± 4.8 | 0.4 ± 3.0 | 11.2 ± 5.5 |
| **Length of stay** | 15.6 ± 27.4 | 15.6 ± 28.4 | 18.8 ± 25.2 |
| **Length of stay > 14 days** | 155 (33.8%) | 127 (33.2%) | 20 (41.7%) |
| **28-day mortality** | 30 (6.6%) | 13 (3.4%) | 16 (33.3%) |
| **MODS** |  |  |  |
| **None** | 319 (69.7%) | 276 (72.1%) | 23 (47.9%) |
| **Any** | 139 (30.3%) | 107 (27.9%) | 25 (52.1%) |
| **Short** | 86 (18.8%) | 70 (18.3%) | 14 (29.2%) |
| **Prolonged** | 53 (11.6%) | 37 (9.7%) | 11 (22.9%) |

Averages are reported as "median ± median absolute deviation".
\*Patients were considered to be in shock if they presented with a base deficit above six.
SBP = Systolic blood pressure. WCC = White cell count. MODS = Multiple organ dysfunction syndrome.

To complement these clinical findings, we employed a controlled porcine model of trauma and haemorrhagic shock to investigate treatment-modulated transcriptomic responses. Following injury and haemorrhage, fifty pigs were randomised to receive one of four treatments: no volume resuscitation (control, n=6), saline (n=8), saline with noradrenaline (n=9), or blood products (n=27), and were followed for eight hours. Serial blood samples were collected pre- and post-injury, and at five intervals following resuscitation, providing a comprehensive temporal dataset. (**Fig. 1**). Blood samples were taken prior to injury and haemorrhage (“pre-injury”), then at 30 minutes after injury (“post-injury”) and thereafter at 90-minute intervals until study completion.

### Trauma shock- and mortality-associated gene expression

RNA-sequencing of whole blood samples from trauma patients at admission revealed extensive transcriptional changes associated with trauma severity and outcomes. Differential expression analysis, adjusted for age and sex, identified 11,825 genes significantly associated with injury severity (ISS), 5,954 with shock and 3,106 with 28-day mortality (5% false discovery rate [FDR]) (**Fig. 2A, S1; Tables S1-3**). There was a high degree of concordance between shock-associated and mortality-associated differential expression, with a strong correlation between observed log fold changes (Pearson’s *r* = 0.90) (**Fig. 2B**), and 93% overlap of mortality-linked genes with shock-associated genes (**Fig. 2C**). Clustering of shock-associated genes revealed a distinct subset of 42 patients (9.2%) with high incidence of shock (58.5%) and mortality (38.1%) (**Fig. 2D**). In the porcine model, these genes effectively segregated pre-and post-haemorrhage samples, providing initial evidence of conserved cross-species injury responses.

**Fig. 2.**
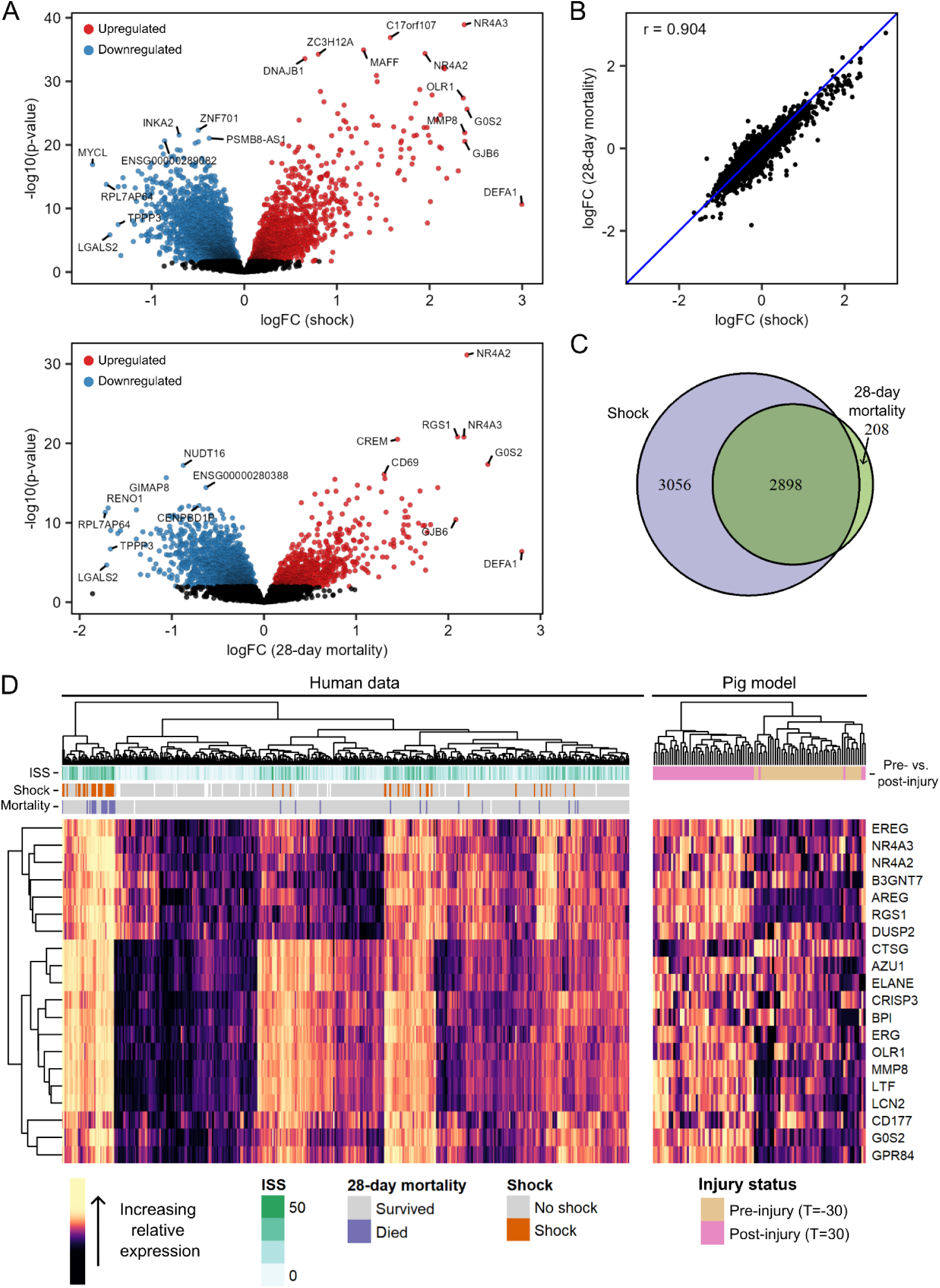
Genes associated with shock and survival in humans. **A**) Volcano plots displaying gene associations with shock (left) and survival (right). logFC = log(Fold Change). **B**) Scatter plot comparing gene log-fold changes for shock (x-axis) and survival (y-axis). Each point represents a gene. Pearson’s r is displayed. **C**) Venn diagram displaying between the genes significantly associated with shock and survival. The purple area indicates shock-associated genes and the green area represents genes associated with survival. **D**) Heatmap for the 30 genes with the greatest log-fold-changes with respect to shock. Note that genes that did not map to expressed porcine genes were excluded for this visualisation (**see Methods**).

Gene Set Enrichment Analysis (GSEA) revealed 196 pathways dysregulated in shock (5% FDR) (**Tables S4-6**), the vast majority of which (98.5%; 193/196) were upregulated in patients in shock. To reduce redundancy and identify major biological themes, we clustered these pathways into 36 distinct functional groups (**Fig. S1; Table S5**). The strongest enrichment was for neutrophil degranulation, characterised by the upregulation of genes encoding neutrophil granule contents (e.g., *DEFA1*, *AZU1*, *MPO*), genes with neutrophil-dominant roles including recruitment (e.g., *CXCL1*, *S100A8*, *S100A12*) and modulators of oxidative stress (e.g., *OLR1*, *HP*, *LTF*). Other strongly upregulated pathways included interleukin and T-cell receptor signaling, natural killer cell and cytotoxic T-cell regulation, and stress-related signalling cascades regulating inflammation, cellular adaptation and tissue repair (including the ATF2, MAPK and oncostatin M pathway terms.)

### Trauma induces conserved transcriptomic changes in humans and the porcine model

To evaluate the translational relevance of our porcine model, we assessed whether human trauma-associated gene networks were preserved across species. Network analysis of the human trauma transcriptome identified 23 distinct co-expression modules, which aligned with the biological processes identified by differential expression in the human cohort (**Fig. S1**). Using a permutation-based approach^17^, we quantified the preservation of these networks in the porcine model, revealing significant conservation of 87% (20/23) of modules (q < 0.05), though with varying strength (**Fig. 3A; Table S7**).

**Fig. 3.**
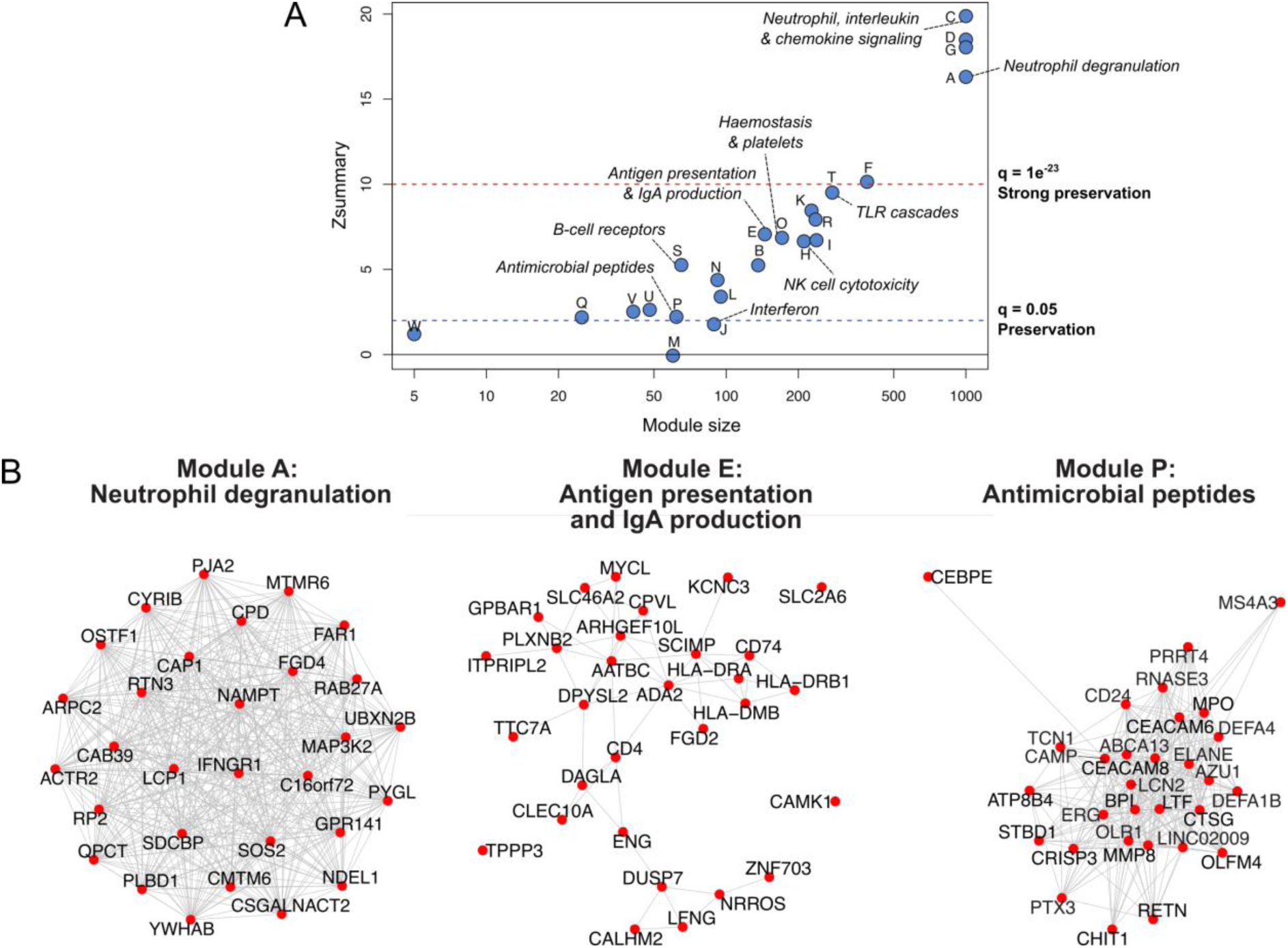
Preservation of human gene coexpression networks in the porcine model. **A**) Preservation of human gene coexpression networks in the porcine dataset. Higher values of Z_summary_ indicate greater preservation. Functional enrichments are shown for selected modules. Modules which had a preservation Z_summary_ score less than 1.96 (q > 0.05) were not considered to be preserved, modules with a Z_summary_ score greater than or equal to 1.96 (or q = 0.05) were considered to be preserved. Modules with a Z_summary_ score greater than or equal to 10 (or q = 1 × 10^−23^) were considered to be highly preserved. **B)** The top genes by module kME for modules A (strongly preserved), E (preserved) and P (borderline preserved). Lines between genes indicate correlation matrix adjacency weights.

Modules governing neutrophil degranulation and chemokine signaling showed remarkable preservation (q < 1×10^−23^) between species (**Fig. 3A, B**). Other key trauma-associated processes displayed moderate to strong (q < 1×10^−5^) preservation, including NK cell cytotoxicity, adaptive immune processes and haemostasis pathways. The robust preservation of inflammatory and immune response modules supports the porcine model’s translational relevance for studying the molecular response to trauma. However, some modules representing immune processes displayed less concordance in the porcine transcriptome. Module P, enriched for antimicrobial peptides (**Fig. 3B**), was weakly preserved, while module J, enriched for interferon signaling pathways (**Table S7**), was considered non-preserved (q > 0.05). These less strongly preserved modules converge on innate antimicrobial defence pathways, which could reflect evolutionary divergence in the transcriptional regulation of specific mechanisms between humans and pigs.

### Transcriptional changes induced by trauma shock are modifiable by therapeutic interventions

Having established that key inflammatory and immune response networks were broadly preserved between human trauma patients and the porcine model, we next used this model to assess the effects of treatment on the trauma transcriptome. Consistent with prior clinical studies^18–22^, survival was different between the volume resuscitation groups, with highest survival with blood product transfusion (27/27), and reduced in those receiving saline only (6/8) or saline with noradrenaline (6/9), and no survival in the untreated control group (**Fig. 4A**).

**Fig. 4.**
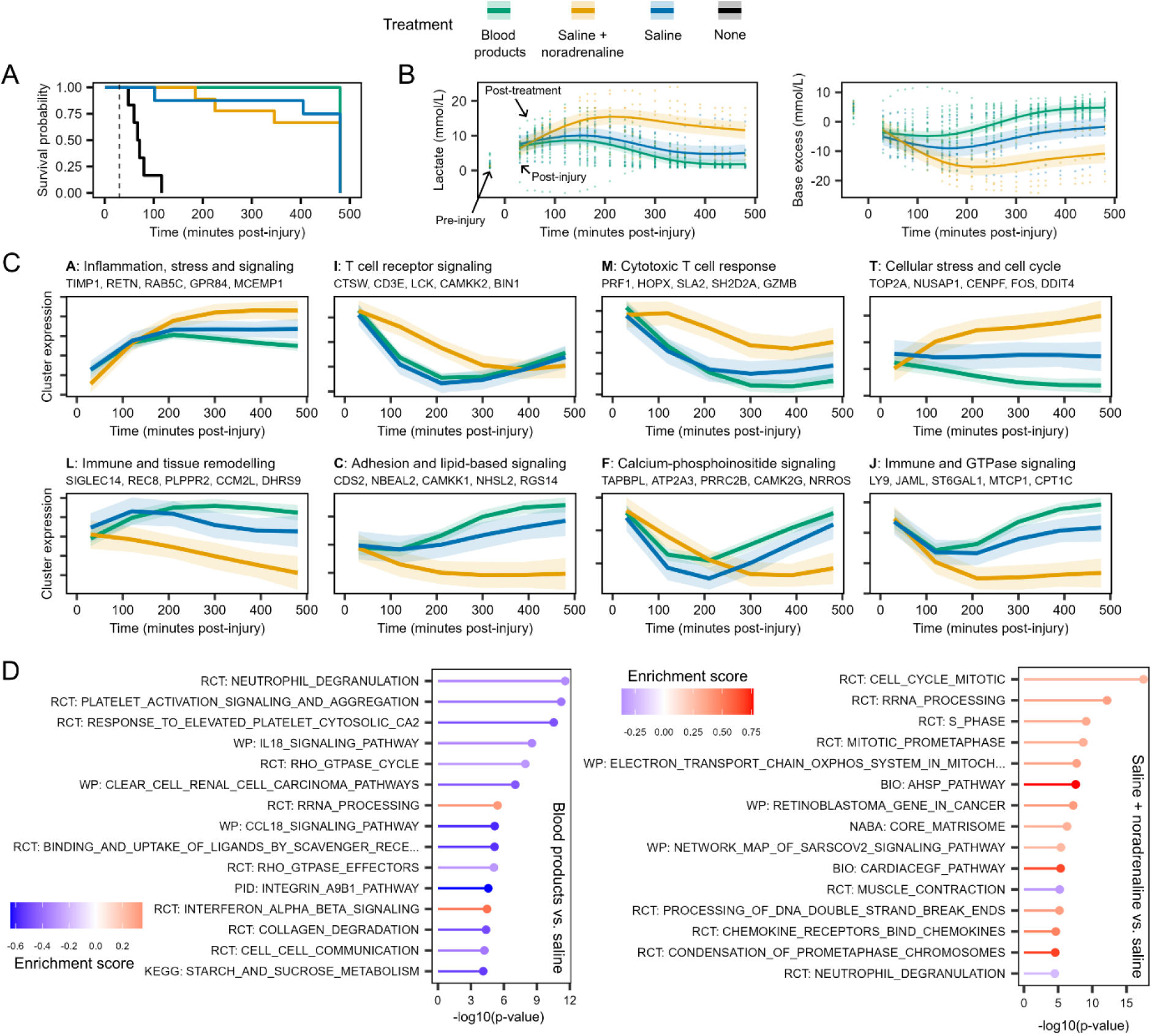
Genes and pathways associated with treatment in a porcine model of trauma. **A**) Survival curves for porcines, stratified by treatment group. The dotted line indicates the post-injury timepoint. **B**) Temporal profiles of lactate (top) and base excess (below) over time in porcines, stratified by treatment group. Lines and shaded areas represent estimated means and 95% confidence intervals, respectively. Points indicate underlying data. The pre-injury (T=-30) samples are plotted but were not included in the modelling process. **C**) Representative expression patterns for clusters of genes with similar temporal profiles. Lines, shaded areas and colour indicate estimated means, 95% confidence intervals and treatment group, respectively. Plot titles indicate the cluster letter, name and five key genes. **D**) The top 15 most significantly enriched pathways for signatures of treatment for the comparison of the “blood products” and “saline” treatment groups (left) and “saline + noradrenaline” and “saline” groups (right). Positive scores (red) indicate upregulation of pathways (i.e., upregulation in the “blood products” group, left, or “noradrenaline”, right), and negative scores (blue) indicate downregulation. Pathways ordered by p-value and coloured by enrichment score.

To understand the molecular basis for these differences, we modelled temporal patterns following trauma to identify parameters whose post-injury trajectories differed significantly between treatment groups (time by treatment interaction; TxT). This approach first revealed distinct temporal profiles in metabolic markers (TxT p < 0.05), with blood product resuscitation reversing markers of ischaemia while noradrenaline exacerbated metabolic derangements (**Fig. 4B**). Applying the same models to transcriptional data identified 983 genes whose temporal expression patterns differed significantly between treatment groups (TxT; 5% FDR) (**Table S8**). Clustering of these genes revealed distinct functional groups (**Tables S9-11**). For example, compared to saline treatment, noradrenaline-treated animals showed elevated expression of genes involved in cell turnover and stress response (clusters A, T, G), T-cell signalling (I) and cytotoxic T-cell activity (M) (**Fig. 4C, S2**). In contrast, these gene clusters showed reduced expression in animals receiving blood products.

While this analysis revealed treatment-specific transcriptional patterns in the porcine model, it did not differentiate between changes relevant to human shock pathophysiology and incidental treatment effects. To address this, we examined the overlap between treatment-responsive genes in our porcine model and shock-associated genes in human trauma patients. 40% (396/983) of the genes associated with treatment in the porcine model were also associated with shock in the human patient cohort (**Fig. S3-4; Tables S12-13**). The genes most significantly associated with both treatment and shock included inflammatory mediators (e.g., *CXCL8*, *CRISP3*, *S100A12*, *OSM*), neutrophil surface and granule components (e.g., *CD177*, *LTF*, *MPO*), and regulators of cellular and oxidative stress (e.g., *OLR1*, *GPR84*, *DDIT4*, *RGS1*) (**Fig. S12**). To systematically evaluate how different treatments affected shock-associated pathways, we compared pathway enrichment patterns between human shock and treatment responses (**Fig. 4D; Table S14**). Blood product resuscitation reversed many shock-associated transcriptional changes (Pearson’s *r* = −0.81 with human shock enrichment scores) (**Fig. 4D**; **Tables S13, S15**). This reversal encompassed key shock-induced pathways including neutrophil degranulation and stress-induced signaling pathways (including OSM and MAPK). In contrast, treatment with noradrenaline and saline tended to enhance shock-induced transcriptional patterns (*r* = 0.29), including IL18, chemokine and T-cell receptor signaling pathways (**Fig. 4D**; **Table S16**), although it did moderately suppress some shock-associated pathways including neutrophil degranulation and matrix metalloproteinase activity.

These findings demonstrate that blood products promote resolution of shock-associated pathways while noradrenaline treatment potentially exacerbates stress responses and perpetuates the inflammatory state. Critically, shock-induced transcriptomic trajectories are not fixed and can be modulated by therapeutic interventions.

### Machine learning identifies robust transcriptional signatures of trauma

While our gene- and pathway-level analysis revealed broad transcriptional changes associated with shock and treatment, individual genes often function as part of coordinated transcriptional programs. To identify these higher-order patterns and prioritise clinically relevant signatures, we applied independent component analysis (ICA) to extract distinct transcriptional signatures, which we refer to as “factors”, from the human cohort (**Tables S17-19**). This approach revealed 40 transcriptionally independent factors, which were evaluated for their association with human clinical parameters and their responses in the porcine model (**Fig. 5A; Tables S20-24**). Many factors showed strong alignment with previously identified shock- and treatment-associated genes (**Fig. S5; Table S25**).

**Fig. 5.**
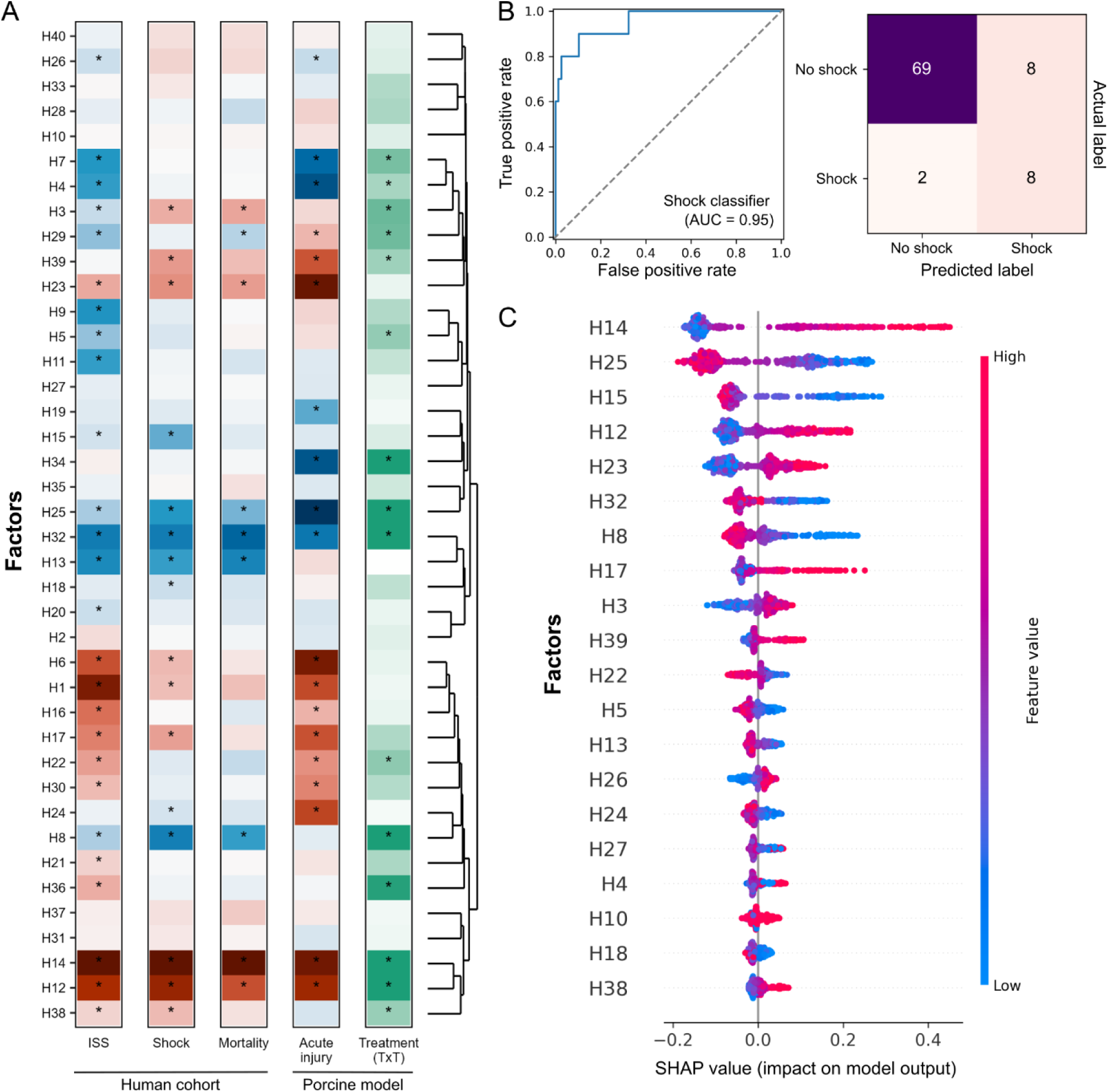
Signatures of human trauma. **A**) Associations of the forty factors derived in humans with both human outcomes and variables in the porcine model. For ISS, shock, survival and acute injury, red indicates upregulation, blue indicates downregulation. For the treatment columns, green indicates significance level. For all columns, asterisks denote significance at 5% FDR. The treatment “interaction” column indicates significance level for the time-by-treatment (TxT) analysis, indicating factors that have dynamic temporal progressions depending on treatment type. Factors are ordered by hierarchical clustering based on their expression. **B**) Receiver operating characteristic (ROC) curve (left) and confusion matrix (right) are shown for the prediction of shock from factor expression using XGBoost. The area under the curve (AUC) is displayed. **C**) SHapley Additive exPlanation plots. Colour indicates the feature value (i.e., high or low gene expression). SHAP values indicate impact on the output, with higher values increasing the likelihood of predicting a patient as having shock. Factors are ordered by their average impact.

To validate these factors, we examined their consistency across both species and independent human cohorts. Of the 26 injury severity-associated signatures, 19 showed concordant responses following trauma in the porcine model (**Fig. 5A**). Analysis of an independent human cohort^9^ demonstrated strong correlation between effect sizes (*r* = 0.88), comparing the estimated effects of ISS in our cohort with the estimated effects of critical injury in the validation cohort, with 22/26 factors showing consistent directionality (**Fig. S6; Tables S26-27**).

To prioritise clinically relevant factors, we developed machine learning models using factor expression signatures as predictive features. Factor models could accurately infer the presence of shock from the factor expression (AUC = 0.95, F1 = 0.62) (**Fig. 5B; Tables S28-29**). We were also able to identify patients with critical injury using the factors (AUC = 0.93; F1 = 0.78) and predict 28-day mortality (AUC = 0.86, F1 = 0.44) (**Fig. S7-8; Tables S28-29**). These models achieved comparable performance to those using gene expression as input directly (**Fig. S8-9; Tables S28-29**). We quantified each factor’s contribution to predictions using SHapley Additive exPlanations (SHAP)^4^, which provided a systematic ranking of each factor’s contribution to outcome prediction (**Fig. 5C, S7-8; Tables S30-35**). These prioritised factors provided a framework for subsequent analyses of treatment effects and potential therapeutic targets.

### Prioritising human shock-induced factors that are responsive to treatment

To identify therapeutically relevant molecular signatures, we considered the set of eight factors that were associated with shock in humans and displayed treatment-dependent temporal profiles in the porcine model (5% FDR; TxT interaction). We termed these “treatment-sensitive shock factors” (TSS factors) and functionally characterised them based on their aligned genes and pathway enrichments (**Fig. 6A; Tables S17, S36-37**).

**Fig. 6.**
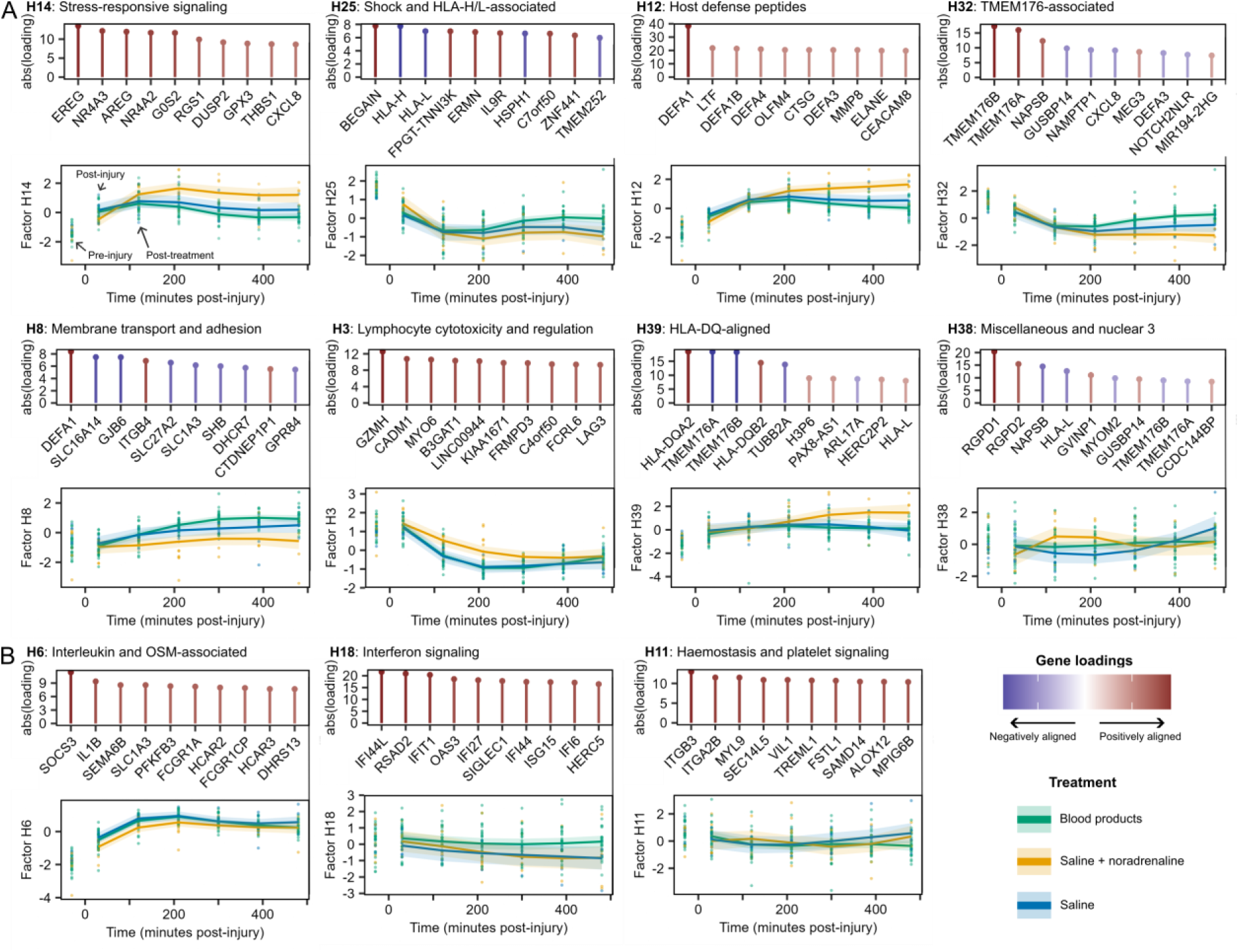
Summaries for key factors. **A/B**) Summary panels for factors, including factor ID and name (**Table factor_summaries**), the top three enriched pathways, the loadings for the most highly aligned genes and the estimated expression of the factor in the porcine model. Temporal expression plots displaying means (lines) and 95% confidence intervals (shaded areas) of factor expression in the porcine data, stratified by treatment group, with points indicating underlying data. **A**) The factors associated with shock and having dynamic temporal profiles depending on treatment group (TxT interaction; 5% FDR), ordered by their impact on shock prediction (Fig. 5C). **B**) Examples of factors that were significantly (5% FDR) associated with human outcomes but not treatment type.

Factor H14, which emerged as the strongest predictor of injury severity, shock and mortality (**Fig. 5C, S7-8**), was upregulated in shock (**Fig. 5A**) and showed sustained elevation following treatment with noradrenaline and saline, when compared to saline alone or blood product resuscitation (**Fig. 6A**). This factor was enriched for MAPK signaling and downstream transcription factors AP1 and ATF2, primarily driven by its alignment with stress-related transcription factors (including *NR4A1-3, FOS*, *FOSB*, and JUN family members), MAPK phosphatases (e.g., *DUSP1*) and the chemokine *CXCL8*. Taken together, these results suggest a role for H14 in coordinating cellular adaptation to trauma.

Several other TSS factors had clear functional links to the immune response to trauma. Factor H12, which displayed similar expression patterns as H14, was enriched for neutrophil degranulation and antimicrobial peptide pathways (**Fig. 6A**). H3, also upregulated in shock, was aligned with IL12 pathway and T-cell activation genes (e.g., *CD8A, CCR5, GZMA*) and markers of cytotoxic lymphocytes (e.g., *KLCR3, FASLG, CADM1*), and correlated with resting NK cell markers (*r* = 0.80) (**Fig. S10; Table S38**). Factor H25, the second strongest shock predictor, was downregulated in shock but elevated by blood product transfusion. It represents heat shock response (e.g., *HSPH1*, *FKBP4*, *HSPA8*) and T-cell receptor signaling (e.g., *TNFRSF9*, *IL1B*, *RIPK2*) pathways that are upregulated in shock.

There were many non-TSS factors that displayed associations with treatment type or human clinical parameters. While we did not prioritise these for follow-up, they could nonetheless represent important disease processes. Some exhibited treatment-dependent temporal profiles without shock association (**Fig. S11**). Others maintained sustained differences between treatment groups but lacked dynamic TxT responses, including factors enriched for neutrophil degranulation (H1), natural killer cell activity (H23) and immunoglobulins (H26) (**Fig. S12**). There were also factors that were not significantly affected by different therapeutic approaches (**Fig. 6B, S13**). These included H6, enriched for interleukin, neutrophil, and oncostatin-M signaling pathways; H11, associated with haemostasis and platelet activation; and H18, aligned with interferon response genes and pathways.

This cross-species analysis prioritised 8 transcriptomic factors that are induced by trauma shock and can be modulated by treatments with known clinical effect. Suppressible pathways include those associated with stress-related pathways, neutrophil activation and cytotoxic lymphocyte actions. Other factors, such as those for haemostasis and interferon responses, did not change with treatment and are therefore potentially less relevant for novel therapies. Overall, these findings provide a framework for identifying modifiable subsets of shock-induced transcriptional changes for selecting future therapeutic candidates.

### Drug discovery identifies candidate trauma shock therapeutics

To identify potential therapeutic interventions which could replicate or augment the beneficial effect of blood component resuscitation on trauma shock, we compared both our eight TSS factors and differential expression profiles (for injury severity, shock and 28-day mortality) with drug-induced expression profiles from the LINCS L1000 project^23^. This analysis revealed 5,725 significant signature-perturbagen associations (5% FDR) (**Fig. S14-15; Table S39**).

Key shock- and survival-predictive factors showed notable therapeutic alignments (**Fig. 7A, S14**). For example, factor H14 was strongly reversed by the anti-inflammatory agent piroxicam, while H12’s signature was counteracted by pentoxifylline, a phosphodiesterase inhibitor known to suppress neutrophil activation^24^. A follow-up analysis considering the drugs that interact with shock- and treatment-associated genes supported these findings, identifying both immunomodulatory compounds that interact with elevated inflammatory mediators (e.g., CXCL8, OSM, IL6R) and PDE4 inhibitors, driven by the upregulation of *PDE4B* and *PDE4D* in shock (**Fig. S16; Table S40**).

**Fig. 7.**
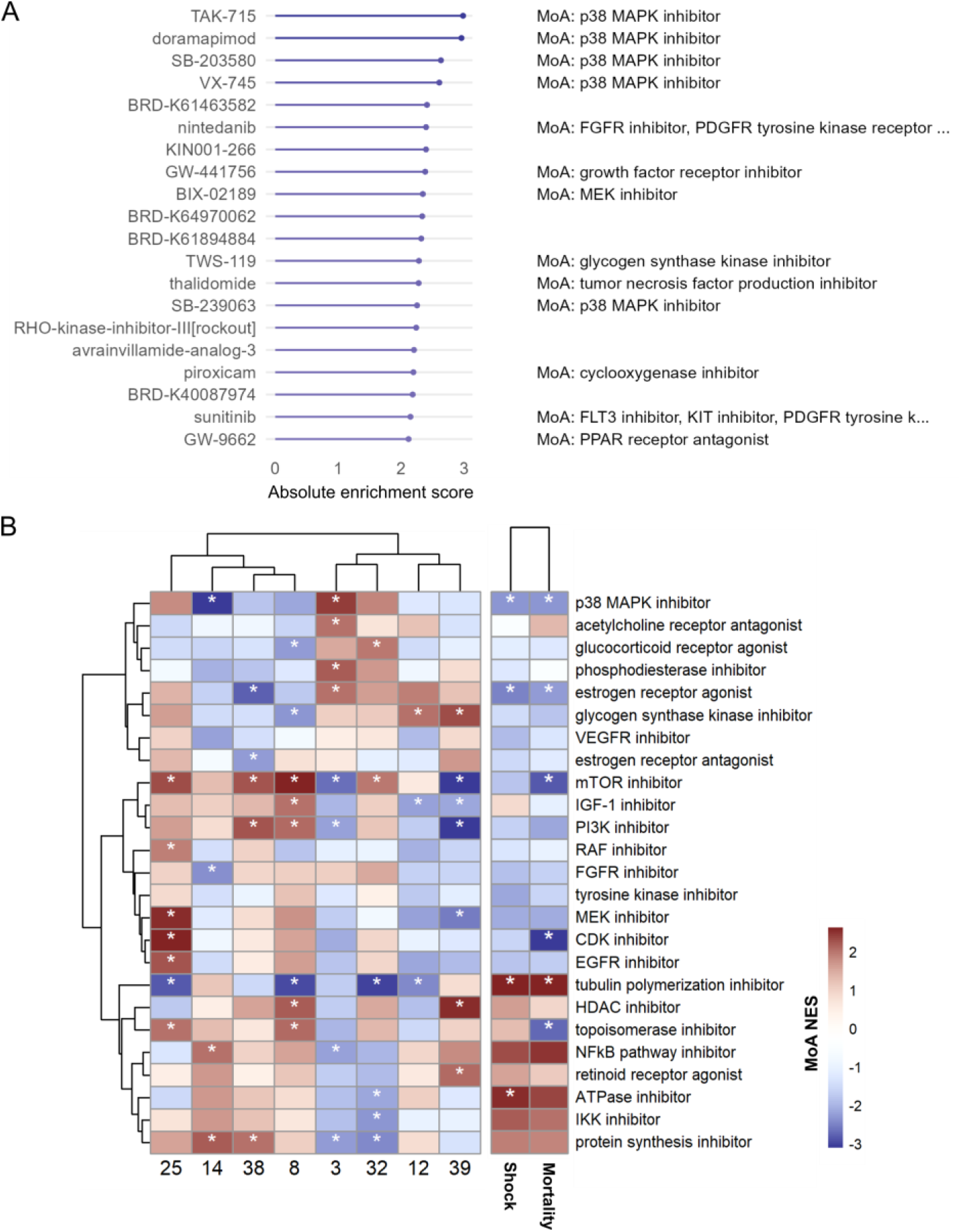
Alignment of factors and outcomes with drug mechanisms. ***A)*** Dotplot indicating enrichment of drug perturbations for factor H14. Negatively aligned perturbations are shown that reverse the H14 signature. The top twenty significantly enriched (5% FDR) perturbations are shown, based on NES score. The MoA(s) and target(s) are shown alongside each perturbation, where available. **B)** Perturbations were grouped by their reported mechanism of action (MoA) and gene set enrichment analysis was used to quantify enrichment. Normalised enrichment scores (NES) are visualised for the top 50 MoAs based on their maximum absolute NES (y-axis) against factors (based on loadings; x-axis, left) and human outcomes (based on differential expression estimates; x-axis, right). Factors are included that were associated with shock in humans and treatment group (main or interaction effect) in the porcine model. Asterisks indicate significance (5% FDR). Rows and columns are ordered by hierarchical clustering of NES scores (based on Ward’s method with Euclidean distance).

To identify broader therapeutic patterns, we next grouped drugs by their mechanisms of action (MoA) and assessed enrichment patterns for shock- and treatment-associated factors (**Fig. 7B; Table S41**). The strongest association was observed between p38 MAPK inhibitors and factor H14. This drug class displayed negative associations with factor H14 and all three clinical outcomes, suggesting these signatures could potentially be normalised by this treatment. The four compounds most strongly reversing H14’s signature were p38 MAPK inhibitors (**Fig. 7B**). This association was mechanistically supported by H14’s enrichments for MAPK-related stress response pathways and downstream transcriptional regulators. The p38 MAPK pathway plays a central role in inflammatory and stress responses^25,26^, making it an attractive target for modulating maladaptive shock-induced changes.

Several other therapeutic classes targeting growth and survival pathways showed potential benefit. mTOR inhibitors were negatively associated with mortality and factor H3, while MEK, EGFR, and CDK inhibitors showed negative associations with shock and survival outcomes alongside positive associations with H25. In contrast, certain drug classes appeared to worsen rather than ameliorate trauma signatures. Tubulin polymerization and protein synthesis inhibitors positively aligned with shock signatures and factor H14, while NFκB pathway inhibitors showed similar pro-inflammatory patterns.

These findings provide a molecular framework for drug repurposing for ischaemia protection in trauma care, with therapeutic candidates targeting specific inflammatory and stress-related pathways.

## Discussion

Haemorrhagic shock in trauma remains an intractable contributor to organ failure and death^2,3^. To our knowledge, this study provides the first largescale human transcriptomic dataset that focuses specifically on trauma haemorrhagic shock and tests the treatment responsiveness of leukocyte transcript programmes within an experimental medicine framework. We found a dramatic overlap (93%) between the global shock transcriptomic response and the 28day mortality gene signature. In the porcine model, noradrenaline resuscitation exacerbated the shock-induced transcriptomic changes, while blood product resuscitation tended to ameliorate the response – mirroring observed clinical and experimental outcomes^18–22,27^. Time-treatment interaction testing across human and porcine models allowed us to identify outcome-critical expression factors that responded to treatment, while other profiles suggested a fixed response to injury. Central to the resuscitation-modifiable shock response were stress-responsive signalling pathways, neutrophil activation and degranulation, and innate lymphoid cell cytotoxicity and regulation. These analyses therefore identify specific modifiable pathways for mechanistic focus and drug discovery.

Understanding the human immune response to injury has been hampered by the massive transcriptomic activation that occurs, often likened to a “genomic storm”^10^. Unlocking the deterministic pathways within this response is key to moving forwards in discovery and translation. Multiple approaches are being explored^9,10^, out remain principally wide-scale and observational, even in experimental studies^28,29^. Here, the randomised intervention model of the animal arm provided a platform to explore treatment responsiveness, as well as an external validation set. Our latent factor approach summarized coordinated biology, reduced the multiple testing burden, and allowed parallel comparison of the human and porcine responses. This model of combining human observational data and experimental randomisation provides a powerful approach to mechanistic discovery.

The transcriptomic factors most strongly linked to shock and most predictive of mortality were also those that were responsive to treatment. Interestingly, some components traditionally thought of as critical to the inflammatory response to trauma, such as interleukin signalling (factor H6) and platelet activation (factor H11), did not predict outcome and were not responsive to treatment. The dominant treatment-responsive shock factor predictive of mortality was the stressresponse module H14. Wholeblood resuscitation suppressed H14, whereas noradrenaline amplified it, consistent with the known effects of these interventions on systemic inflammation and outcomes^18,22^. H14 was enriched for p38mitogenactivated protein kinase (MAPK) and activator protein-1 (AP-1) signalling. The p38-MAPK system is known to amplify cellular stress by stabilising transcripts that encode pro-inflammatory cytokines and by activating AP-1–dependent genes that drive neutrophil oxidative burst, endothelial permeability and metabolic suppression^30,31^. Sustained p38-MAPK activation precedes mitochondrial dysfunction and microvascular thrombosis in rodent haemorrhagic shock^28^, processes closely associated with fatal multiple-organ dysfunction. In the LINCS screen, p38 inhibitors produced the strongest inverse connectivity scores against H14. Previous rodent models of trauma haemorrhage showed pharmacological blockade of p38MAPK with SB203580 or related compounds limited renal, hepatic and cardiovascular dysfunction and reduced inflammatory cytokine production^32–35^. This agreement between human and porcine transcript shifts, *in silico* predictions and experimental investigation provides causal support for our methodology. Several selective p38 inhibitors identified by our drug–signature analysis, including TAK-715, doramapimod, SB-203580 and VX-745, have established pharmacology, acceptable safety profiles, and Phase II human experience, underscoring translational feasibility^36^. Notably, another molecule in this class, dilmapimod (SB-681323) was evaluated in a Phase IIa trauma cohort at risk for ARDS, where it proved safe and modulated inflammation^37^. Although development of p38 inhibitors has been hampered by issues with chronic administration in inflammatory disease^38^, trauma represents a distinct, time-limited therapeutic window in which brief pathway suppression is both mechanistically justified and operationally viable. These precedents support our deployment of this experimental medicine framework on a clinical trauma platform with integrated transcriptomic and physiological profiling^39^, enabling a practical bridge between discovery and early human proof-of-concept.

Other treatment responsive factors included H25, the next strongest mortality-associated factor, which captured a heat-shock response. Expression of H25 fell after shock but was partially restored by whole-blood transfusion, with no appreciable change under noradrenaline. This may suggest that early heat shock stress is amenable to correction when oxygen-carrying capacity and buffering are promptly re-established. Factors H12, a neutrophil-granule programme, and H3, an NK/CD8 cytotoxic module, rose sharply after shock. Noradrenaline increased expression further during resuscitation, while blood component resuscitation maintained or normalised their responses. Conversely, Factor H8, the fourth-strongest determinant of mortality in the multivariable model, was dominated by membrane-transport and antimicrobial genes. Blood resuscitation partially restored its expression whereas noradrenaline further suppressed it, suggesting an early barrier-defence deficit that is sensitive to resuscitation strategy. Collectively, these treatment-responsive factors expose discrete neutrophil, cytotoxic-lymphocyte, heat-shock and barrier defence nodes that converge on stress-kinase signalling, delineating tractable molecular entry points for future mechanistic investigation.

Virtual screening using transcriptomic signatures has shown promise across diverse therapeutic areas^40–44^, though these studies typically rely on observational data without experimental validation of treatment effects. While porcine gene expression has been assessed previously in trauma contexts^45–47^, these studies did not perform treatment interventions. Our data provided a unique opportunity to study treatment-responsive pathways. However, our analytical approaches rely on the identification of robust gene signatures and the relevance of the porcine transcriptomic response for human trauma. To this end, we tested our factors in external datasets, finding remarkable concordance (r = 0.88), despite differences in patient selection and transcriptomic platform. Furthermore, we found that core human trauma gene co-expression networks displayed strong preservation in the porcine model. However, there were notable exceptions, including interferon and antimicrobial peptide pathways, which could reflect species-specific immune architecture^48,49^. Whilst these findings provide confidence in our results, they also highlight the importance of pathway-specific considerations when translating findings from porcine models to human trauma care.

Our study is limited in several ways. First, by focusing on the white cell transcriptome, we may have missed key trauma responses that occur primarily in the endothelium and subendothelial tissues. Furthermore, bulk whole-blood analysis can suggest but not confirm cell-specific responses, but single-cell methods are required achieve such resolution. Similarly, our drug repurposing analyses relied on the assumption that perturbation signatures from L1000 cell lines, which are not derived from tissues directly involved in the trauma response, can be meaningfully compared to trauma signatures *in vivo*. Additionally, L1000 platform’s limited gene coverage may have missed important trauma-related genes^23^. Cross-species comparisons were constrained both by differences in sampling time points between human and porcine samples and by the use of Ensembl homologs for gene mapping, as incomplete or imperfect homology may reduce the accuracy of these analyses. Finally, despite leveraging an experimental model, our findings remain associative and specific mechanistic studies are required to explore the involvement and relative dominance of these pathways in developing tissue injury, organ dysfunction and death. A human experimental medicine study that applies defined pharmacological interventions and tracks temporal transcriptomic changes would be a valuable next step.

In summary, our cross-species experimental medicine framework identifies tractable, treatment-responsive components of the trauma shock response and highlights p38-MAPK signalling as a dominant, modifiable driver of maladaptive inflammation and cellular stress. Multiple selective p38 inhibitors with established human safety data, including those already evaluated in critical-illness and trauma-adjacent contexts, demonstrate that transient systemic p38 inhibition is clinically feasible. These precedents, together with the mechanistic prioritisation presented here, provide an immediate translational pathway for prospective evaluation of p38-MAPK inhibition within controlled trauma platforms, More broadly, we provide a systematic map linking trauma-induced molecular programmes to perturbation strategies, offering a resource to guide targeted therapeutic development, accessible via our interactive web portal (https://shiny-whri-c4tb.hpc.qmul.ac.uk/traumashiny/).

## Materials and Methods

### Human patient cohort

Patients were recruited between February 2008 and March 2019 through the Activation of Coagulation and Inflammation in Trauma (ACIT-II) study^16^, a prospective platform cohort study evaluating aspects of coagulation and inflammation in trauma patients (NHS REC: 07/Q0603/29). Patients at the Royal London Hospital were enrolled into ACIT-II in the emergency department. Patients were eligible for enrolment given that they were admitted within two hours from injury. Admission blood samples and physiology data were obtained on arrival, and further clinical data were collected daily over the following 28-days or until discharge from hospital or death. The Injury Severity Score (ISS) for each patient was calculated once all injuries had been fully assessed and documented. Patients were selected to represent a range of injury severities: low (ISS 0-3; n=80), mild (ISS 4-8; n=80), moderate (ISS 9-15; n=81), severe (ISS 16-24; n=79) or critical (ISS >24; n=138) (**Table 1**). Only patients with blunt trauma were included, while those with penetrating injuries or Traumatic Brain Injuries (TBI) were excluded. Patients with a base deficit greater than six were defined as being in early physiological shock.

### Porcine model of traumatic haemorrhagic shock

This was a randomised study in terminally anaesthetised female cross-bred Large White pigs; the study underwent local ethical review and was carried out under the authority of Animals (Scientific Procedures Act 1986. The full detailed experimental protocol has been previously published^14^); the model is described in brief below.

Initially, animals were anaesthetised and instrumented for continuous physiological monitoring and sampling; blood samples were collected upon initiation of the surgery (line-in, T = −150 minutes). Following surgery, animals’ physiology was allowed to recover for 60 minutes prior to pre-injury (T= −30 minutes) samples being taken. The injury phase comprised three components, a soft tissue injury created using a captive bolt (4 shots to the right thigh), controlled haemorrhage (30% blood volume), and a snare around the liver lobe was pulled to initiate an uncontrolled haemorrhage. Animals then entered a 30 minute “buddy aid” phase after which samples were collected (post-injury; T = 30 minutes). Animals were then treated with either saline alone (n = 8), saline and noradrenaline (n = 9), or blood products (n = 27). The blood group is comprised of animals treated either with fresh whole blood (FWB; n = 9), fresh frozen plasma (FFP; n = 9) or packed red blood cells (PRBC) and FFP in a 1:1 ratio (n=9). For the purposes of downstream analysis, the animals administered FWB, FFP or PRBG:FFP were combined to form a single “blood products” group. Blood products were collected and prepared from donor animals as previously described^50^ in accordance with UK donation guidelines. Prior to use, all donor products were forward and reverse matched to recipient blood. Blood samples were collected throughout the treatment phase (T=120, 210, 300, 390 and 480 minutes). At the end of the experiment (480 minutes following the start of treatment), or when the SBP <10mmHg, animals were culled with an overdose of pentobarbitone (Euthatal, Merial Animal Health Ltd, Harlow, UK).

For transcriptomic analysis, blood samples (100 µL) were collected into RNAprotect® Animal Blood Tubes (Qiagen), incubated overnight at room temperature before storage at −35°C.

### RNA sequencing protocol

Total RNA was extracted using the PAXGene RNA extraction kit (Qiagen, Hilden, Germany) according to the manufacturer’s instructions, with a DNase step included to remove contaminating DNA. RNA samples were assessed for quantity and integrity using the NanoDrop 8000 spectrophotometer v2.0 (ThermoScientific, USA) and Agilent 2100 Bioanalyser (Agilent Technologies, Waldbronn, Germany), respectively. All samples that displayed an RIN score of 6.5 or higher were used for RNA library preparation (NEBNext Globin & rRNA Depletion Kit). Samples were randomized before library preparation and 60bp paired-end reads were generated for each library using NextSeq2000 P3 100-cycle kit (Illumina Inc., Cambridge, UK).

For the human data, following quality control, gene counts were calculated using Salmon^51^ in combination with the Gencode v44 GRCh38 assembly and annotations. Quantification of the porcine transcripts was achieved with the kallisto pseudoaligner^52^ in combination with the Ensembl Sus scrofa reference (release 110). For both datasets, the raw count data were imported with tximport^53^ in R. Normalisation factors were calculated using edgeR’s “Trimmed Mean of M-values”^54^, based on the approach recommended in the tximport vignette^53^. Except for differential expression analysis, we converted the data to log-transformed counts per million (LogCPM) for downstream use.

Further details are available in the **Supplementary Methods**.

### Statistical analysis

Differential expression analysis was performed to assess gene-level associations using the (dream) pipeline^55^ implemented in the variancePartition package^56^. For the human cohort, models were adjusted for age and sex. For the porcine model, random intercepts were added to take into account repeated measurements. Two-sided tests were used in all instances. Multiple testing correction was carried out using the Benjamini-Hochberg procedure to control the false discovery rate at 5%. Functional enrichment analysis was carried out using the Gene Set Enrichment Analysis (GSEA) procedure, implemented in the fgsea package^57^. Pathways were defined using the Molecular Signatures Database (MSigDB) C2 canonical pathways^58^, excluding the Chemical and Genetic Perturbations (CGP) subcategory. To reduce redundancy in these pathways, we used the SimplifyEnrichment package^59^ to cluster similar pathways based on shared genes.

To facilitate multi-gene analyses, we developed the “ReducedExperiment” framework for identifying, manipulating and analysing transcriptomic factors, implemented in R and available from Bioconductor (https://bioconductor.org/packages/ReducedExperiment)^60^. Specifically, within this package we implemented the stabilised ICA algorithm, which allows for the estimation of factor stability and has been shown to enhance reproducibility^61,62^. ReducedExperiment provides helper methods for fitting linear models in a factor-wise manner. We applied these to fit models equivalent to the gene-level differential expression analysis, using R’s^63^ lm function in combination with the car package’s^64^ Anova function. The lmerTest^65^ package was used to fit linear mixed models (LMMs) where random intercepts were necessary. Treatment-by-time interaction (TxT) effects were assed with an LMM with a natural cubic spline to model expression changes over time.

The WGCNA package^17,66^ was used to define groups of coexpressed genes and evaluate their cross-species preservation. The machine learning models were implemented through the scikit-learn^67^ and XGBoost^68^ packages in Python. We randomly partitioned patients into training (80%) and held-out test (20%) sets. We performed hyperparameter optimisation with the Optuna library^69^, maximising the average area under the curve estimated during stratified 4-fold cross-validation in the training set. We quantified the impact of inputs on predictions using the SHAP package^70^. We used the procedure described by the metaLINCS^71^ R package to identify LINCS L1000^23^ small molecule perturbation signatures that could counteract trauma-associated transcriptional changes, focussing on a subset of 20,220 perturbagens corresponding to compounds in the Broad Institute’s Drug Repurposing Hub^72^.

These procedures are described in more detail in the **Supplementary Methods**.

### Interactive portal

We developed an interactive shiny portal (https://shiny-whri-c4tb.hpc.qmul.ac.uk/traumashiny/) to facilitate exploration of the study findings. This resource enables users to: (i) query gene- and pathway-level results across both human and porcine datasets, (ii) sort factors by predictive impact on different outcomes, and (iii) examine detailed factor-specific information including clinical associations, expression patterns, pathway enrichments and drug connectivity analyses. The portal provides a comprehensive interface for investigating trauma-associated transcriptional signatures and their therapeutic implications.

## List of Supplementary Materials

**The supplementary PDF file contains**:

Supplementary Results

Supplementary Methods

Fig. S1 to S15

**Other Supplementary Material for this manuscript includes**:

Tables S1 to S49

## Acknowledgements

This research utilised Queen Mary’s Apocrita HPC facility, supported by QMUL Research-IT. http://doi.org/10.5281/zenodo.438045. We would also like to thank members of the Combat Casualty Care project team at Dstl for their expert technical support of the animal studies presented in this manuscript.

## Author contributions

J.G. and I.W. analysed the data. J.G., R.P., I.W., S.W., E.K., M.R.B. and K.B. contributed to data interpretation. J.G., M.R.B. and K.B. wrote the manuscript. S.W., E.K., M.R.B. and K.B. designed the study. All authors critically reviewed and approved the final manuscript.

## Data and materials availability

We also provide the counts and normalised expression data for both the human cohort and the porcine models as supplementary data files. Additionally, on ArrayExpress we deposited the porcine model reads (E-MTAB-16011) and human cohort counts (E-MTAB-16212). The normalised microarray data for the independent human cohort was obtained from ArrayExpress (E-MTAB-5882).

The code for identifying factors and downstream analysis is implemented in the ReducedExperiment package (https://bioconductor.org/packages/ReducedExperiment)^60^. We also provide notebooks for replicating major analyses on GitHub (https://github.com/jackgisby/trauma_treatment_code).

Finally, we provide an interactive R shiny portal (https://shiny-whri-c4tb.hpc.qmul.ac.uk/traumashiny/; user: traumashiny; password: murky shield flood) that allows users to explore results for individual genes and factors.

## Supplementary Results

### Differential expression (porcine model): Line-in vs. pre-injury

The “line-in” sample was taken near the start of surgery, whereas the “pre-injury” sample was taken prior to injury (**Fig. 1**). The former reflects the effects of the initial surgery, whereas the latter describes the state of the transcriptome after a rest period. We performed differential expression analysis and gene set enrichment analysis to compare these timepoints (**Tables S42-43**).

### Differential expression (porcine model): The effect of injury

We compared the pre- and post-injury samples, to characterise the effect of acute injury (**Table S44**). Many of the pathways enriched for acute injury in the porcine model (**Table S45**) were also observed for injury severity in the human cohort (**Fig. S1**).

### Differential expression (porcine model): Base excess and lactate

We attempted to perform an analysis in the porcine model that was equivalent to the differential expression of the shock outcome in the human cohort (**Fig. 2A, D; Table S2**). We considered associations with base excess and lactate at the post-injury timepoint in the porcine model. We identified far fewer significant (5% FDR) genes (81 for base excess and 54 for lactate) (**Tables S46-49**). In part, this was because there were only 49 samples at the post-injury timepoint, compared to the 458 patients included in the analysis of human shock. Furthermore, at this point in the study, all the pigs had experienced the same controlled conditions, whereas the injuries received by the human patients varied greatly in their severity.

### Shock-treatment genes

There were 396 genes that were both associated with shock and exhibited treatment-dependent temporal profiles (TxT; 5% FDR). Using robust rank aggregation (RRA), we prioritised genes based on their combined strength of association with shock and treatment response (**Fig. S3-4; Tables S12-13**).

We explored therapeutic implications by querying the Drug-Gene Interaction Database (DGIdb) for compounds interacting with the proteins encoded by these genes (**Fig. S3C**, **S16; Table S40**). The analysis revealed several classes of potentially relevant drugs. Immunomodulatory and anti-inflammatory agents, including tocilizumab, cyclosporine, tolmetin and amlexanox, emerged as prominent candidates. PDE4 inhibitors, such as ibudilast and roflumilast, were identified due to the upregulation of both PDE4B and PDE4D in both patients suffering from shock and noradrenaline-treated animals.

### Supervised learning

The expression models marginally outperformed those based on factors for the shock (AUC of 0.97 and 0.95 for genes and factors, respectively) and 28-day survival outcomes (AUC of 0.87 and 0.86 for genes and factors, respectively) (**Table S28**). The gene-based shock and survival models had excellent specificity (1 – false positive rate; shock = 0.90, survival = 0.91) and good sensitivity (false negative rate; shock = 0.80, survival = 0.67), but relatively poor precision (shock = 0.50, survival = 0.33). While these models overpredicted the positive class, the majority of the false positives had high ISS (**Fig. S9D**). Furthermore, we note that most of the patients misclassified as having shock had above average base deficit (7/8), but did not quite reach the cutoff for being defined as being in shock using in this study (base deficit > 6). These findings suggest that the false positives identified by these models likely represent patients that were molecularly severe, and likely had high risk for experiencing poor outcomes. To validate the genes with the highest SHAP values for prediction of 28-day mortality, we constructed survival curves stratified by the expression of these genes (**Fig. S9C, E**).

## Supplementary Methods

### RNA-seq processing

Initial quality control and alignment was performed for the human cohort using the nf-core/rnaseq^1^ nextflow^2^ pipeline (v3.12.0). We evaluated the reads with FastQC^3^ and performed trimming with Trimgalore^4^. One sample fell below the minimum trimmed reads threshold (10,000) and was excluded. Gene counts were calculated using Salmon^5^ in combination with the Gencode v44 GRCh38 assembly and annotations. The porcine data were also evaluated using FastQC^3^. Gene count quantification was carried out with the kallisto pseudoaligner^6^ in combination with the Ensembl Sus scrofa reference (release 110).

The raw count data were imported with tximport^7^. For the porcine counts, we used AnnotationDbi^8^ to map transcript names to gene IDs, whereas for the human cohort we used the mapping created by the Nextflow pipeline. We calculated normalisation factors based on the approach recommended in the tximport vignette^7^. This approach combined per-observation scaling factors for length with the effective library sizes calculated using edgeR’s^9^ calcNormFactors function (method=”TMM”). To filter genes with low counts, we used edgeR’s filterByExpr function. We applied the normalisation factors, converted the data to counts per million (CPM) and log-transformed the data for downstream analysis. For the purposes of differential expression analysis, we instead converted the data to a DGEList object and reapplied calcNormFactors. As a quality control step, we applied principal components analysis to the normalised data, and visualised the samples using scores plots. Based on these, we removed three samples from the porcine data that were extreme outliers.

### Gene annotations

We used the biomaRt package^10^ to map Ensembl IDs to other identifiers. We mapped the human dataset’s genes using the “hsapiens_gene_ensembl” dataset^11^ (v110) to HGNC symbols^12^ and Entrez IDs. We primarily used Ensembl identifiers, however for plots we report the HGNC gene names where available. Likewise, we obtained HGNC symbols and Entrez IDs for the porcine data using the “sscrofa_gene_ensembl” dataset. Additionally, we mapped the Sus scrofa Ensembl IDs to their human homologs, based on the “hsapiens_homolog_ensembl_gene” attribute. For downstream analyses, we filtered out porcine genes that did not have a recorded human homolog.

We used an external microarray dataset from an independent human cohort (E-MTAB-5882). We mapped probes from this data to our human RNA-seq data based on gene symbols.

### Gene expression modelling

Gene-level differential expression analysis was performed using the differential expression for repeated measures (dream) pipeline^13^ implemented by the variancePartition package^14^. Dream permits the incorporation of gene-level random effects to account for repeated measurements. Statistical significance was assessed using empirical Bayes moderated t-statistics. For all differential expression analyses, p-values were adjusted for multiple testing using the Benjamini-Hochberg procedure controlling the false discovery rate at 5%. This pipeline was applied for all gene-level analyses, including human outcome associations, porcine timepoint comparisons, and treatment-dependent temporal trajectories. Associated pathways were identified using Gene Set Enrichment Analysis (GSEA), with the exception of the treatment-by-time interaction models, to which we applied overrepresentation analysis (**see Methods – Pathway enrichment analysis**).

We also fit linear models for clinical measurements (including lactate, base excess and blood parameters) and factors (**see Methods – Factor analysis with ReducedExperiment**). For standard analyses, we employed type II analysis of variance using the car Anova function^15^ on linear models constructed using R’s^16^ lm function. For model formulations that required random effects, we implemented linear mixed models (LMMs) using the lmerTest package^17^. These models were fitted using restricted maximum likelihood estimation (REML), with significance assessed through F-tests incorporating Satterthwaite’s approximation for degrees of freedom^17^.

### Model formulation: Human outcomes

To identify genes and factors associated with clinical outcomes, we modelled expression (“E”) as a function of the outcome of interest. For injury severity, we used log-transformed ISS as a continuous predictor. Shock status (defined as base deficit > 6) and 28-day mortality were included as binary variables. All models included age (continuous) and sex (binary) as covariates. The regression model for these analyses in Wilkinson-style notation was:

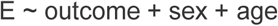

### Model formulation: Porcine timepoint comparisons

To characterise the response to instrumentation and injury, we performed paired comparisons between successive timepoints in the porcine data. The initial response to surgical instrumentation was assessed by comparing line-in (T = −150 minutes) to pre-injury (T = −30 minutes) samples. The acute effect of trauma was evaluated by comparing pre-injury (T = −30 minutes) to post-injury (T = 30 minutes) samples. To account for repeated measurements from the same animals, we included a random intercept term to estimate between-subject variability. The model was:

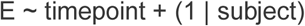

### Model formulation: Clinical associations in porcine model

To identify early markers of physiological deterioration, we considered associations with base excess and lactate (“clinical_parameter”) at the post-injury timepoint (T = 30 minutes). The model formulation was:

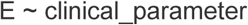

### Model formulation: Treatment-dependent temporal trajectories

We examined longitudinal trajectories of genes, factors and clinical measurements (“variable”) in the porcine model from the post-injury timepoint (T = 30) through study completion (T = 480). To capture non-linear temporal patterns while maintaining model parsimony, we modelled time using natural cubic splines (ns function, splines package in R). Model selection based on Akaike information criterion (AIC) supported using splines with three degrees of freedom (mean AIC=545) compared to more constrained models (AIC=590 and 550 for df=1 and df=2, respectively) or modelling time as an unordered factor (AIC=561). To test whether variables exhibited treatment-specific temporal patterns, we included treatment type as a covariate and an interaction term between time from injury and treatment (TxT). The model formulation was:

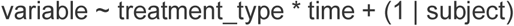

A significant treatment-by-time interaction (TxT) indicated that the temporal evolution of the dependent variable differed between treatment groups. In the absence of a significant interaction term, a significant main effect of treatment without an interaction effect indicated a consistent difference between treatment groups across all timepoints. Finally, a significant time effect alone indicated that the variable changed over time, but followed similar temporal patterns across all treatment groups.

In addition to reporting the anova outputs based on these models, we extracted and plotted predicted means and confidence intervals using the predict_response function from the ggeffects package^18^.

### Model formulation: Independent human cohort

We modelled the expression of factors in an independent human cohort (E-MTAB-5882). Samples were available in the hyperacute window (within 2 hours of injury; “0h”), in addition to 24h and 72h post-injury. Based on their injury severity score, the 39 patients were classified as mild (ISS ≤ 4), moderate (9 ≤ ISS ≤ 13) or critical (ISS ≥ 25). We modelled both time post-injury (“time”) and severity group (“group”) as categorical variables, including their interaction to capture severity-specific temporal patterns. We modelled both the main effects of time and severity, and their interaction. We included both age (continuous) and sex (binary) as covariates. We accounted for batch effects and repeated measurements from the same individual using random intercepts. The model formulation was:

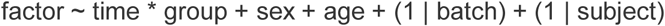

Means and confidence intervals were calculated using the predict_response function from the ggeffects package^18^. To assess the effect of injury during the hyperacute window, we performed post-hoc analysis comparing expression between critical and mild groups at 0h using the emmeans package^19^.

### Clustering temporal profiles

Genes displaying significant treatment-by-time interactions (TxT; FDR < 0.05) in the porcine model were clustered based on their predicted temporal profiles, extracted for the fitted models using the ggeffects package^18^. We clustered genes by average-linkage hierarchical clustering based on correlation distance (1 – Pearson correlation) between their temporal profiles. We defined clusters using the dynamic tree cutting algorithm (dynamicTreeCut R package^20^, deepSplit = 2) with a minimum cluster size of 15 genes.

For each resulting cluster, we calculated a representative expression profile (“eigengene”) based on the first principal component of the cluster genes. We identified hub genes within each cluster by calculating the squared correlation (R^2^) between the gene’s expression and its corresponding eigengene. We identified enriched pathways for each cluster using an overrepresentation test, with the full set of matched human orthologues as the gene universe. We assigned descriptive names to each cluster based on their enriched pathways and hub genes.

### Treatment signature comparisons

We performed pairwise comparisons between the porcine treatment groups for all genes in the porcine model. We calculated predicted temporal profiles for the fitted treatment-by-time models (TxT) using the ggeffects package^18^. For each gene, we took the difference between the estimates for each pairwise comparison at each timepoint. We then defined an aggregated signature based on the maximal difference across the timepoints. We then identified pathways by applying (GSEA) to the signatures.

### Prioritising genes associated with shock and treatment

To identify genes both strongly associated with human shock and exhibiting distinct treatment-dependent temporal profiles, we used robust rank aggregation^21^ (RRA). This method identifies features that are ranked higher than expected across multiple lists. We integrated two measures as input: the absolute log-fold change for shock association and the log-transformed p-values for the treatment-by-time interaction term. RRA generates a significance score analogous to a *P*-value. We applied a −log10 transformation to these values, so that a larger score indicates stronger associations across both shock and treatment. We report RRA scores for genes reaching significance (5% FDR) in both analyses.

### Identifying overrepresented gene-drug interactions

We leveraged the Drug Gene Interaction Database (DGIdb latest, accessed 11/07/2024) to identify potential therapeutic targets among genes associated with both human shock and treatment response (TxT) in the porcine model (5% FDR). For each drug, we performed Fisher’s exact tests to assess whether its known gene interactions were overrepresented among our identified genes compared to the background of all tested genes. We considered drugs with at least two target genes and controlled for multiple testing using the Benjamini-Hochberg procedure (5% FDR). We filtered out generic drug categories from the final results (e.g., “IMMUNOTHERAPEUTIC AGENT”, “THERAPEUTIC CORTICOSTEROID”) but we did not filter interactions based on their type, score or source.

### Factor analysis with ReducedExperiment

We applied independent component analysis (ICA) to identify distinct transcriptomic factors. We used the stabilised ICA (sICA) algorithm, which involves running multiple ICA iterations to provide an estimate of factor stability and enhance reliability^22^. A systematic comparison of dimensionality reduction approaches demonstrated superior inter-dataset reproducibility of stabilised components in transcriptomics data^23^. We used the “Most Stable Transcriptome Dimension” (MSTD) protocol^24^ to determine the optimal number of components, yielding 40 distinct factors in our human cohort. We used the sICA and MSTD implementations in the ReducedExperiment Bioconductor package^25^, and also used this package for downstream analysis, described below.

To identify genes associated with each factor, we analysed scaled loadings using the package’s getAlignedGenes method. Genes were classified as highly aligned with a factor if they met two criteria: the loading magnitude had to exceed half of the factor’s maximal loading, and the loading had to rank within the top 1% for that factor (**Table S36**). To provide a biological annotation for factors, we performed GSEA-based enrichment analysis on the factor loadings using the ReducedExperiment runEnrich function, with 10,000 permutations and a convergence threshold of 1×10^−50^.

Projection of factors into additional datasets was accomplished using the ReducedExperiment projectData function. Sample-level scores were calculated using factor loadings and normalised transcriptomic data from the target dataset. We performed cross-species and cross-platform analyses (porcine cohort and external microarray data, respectively), restricting these analyses to genes measured in both datasets.

To evaluate statistical associations between factors and sample-level variables, we used the associate_components function.

### Immune cell deconvolution

We used CIBERSORTx^26^ to predict the cell fractions present in the human blood samples. Imputation was performed based on the normalised RNA-seq data for 22 classes of immune cell using the LM22 signatures^27^. The application was run with batch correction enabled (B-mode) and quantile normalisation disabled.

### Comparing signatures to gene-drug perturbations

To identify potential therapeutic interventions, we leveraged the LINCS L1000 dataset^28^ to search for drug-induced gene expression signatures that could counteract trauma-associated transcriptional changes. We analysed a subset of 20,220 perturbagens corresponding to compounds in the Broad Institute’s Drug Repurposing Hub^29^, allowing us to focus on drugs with established clinical relevance (approved or tested in human trials).

We implemented a multi-level analytical pipeline based on the metaLINCS R package^30^ to evaluate both the distinct ICA-derived factors and differential expression signatures for clinical outcomes (ISS, shock, 28-day mortality). This approach offers advantages over conventional single signature matching by enabling systematic aggregation at both the perturbagen and mechanism of action (MoA) levels.

The analysis involved three stages:

1. For each trauma signature, rank correlations were calculated with L1000 perturbation profiles, using standardised expression values (level 5 z-scores).
2. GSEA was employed to identify perturbagen-level enrichments, aggregating results across different experimental conditions (cell lines, doses and time points) for each compound. We filtered out perturbagens that had fewer than five instances per perturbagen.
3. Finally, a second GSEA step was performed on the perturbagen-level enrichment scores grouped by MoA classifications from the Drug Repurposing Hub. This additional layer of analysis enabled the identification of drug classes and broader therapeutic mechanisms associated with trauma signatures. MoA groups were removed that mapped to fewer than three compounds.

GSEA was calculated with the fgseaSimple function implemented in R’s fgsea package^31^ using 100,000 permutations. We adjusted p-values using Benjamini-Hochberg to control the false discovery rate at a level of 5%.

We applied two approaches to prioritise robust therapeutic associations. Firstly, we focused on factors that were both associated with human shock and showed treatment-dependent temporal changes in our porcine model (FDR < 0.05). Secondly, we confirmed that perturbagens and MoAs of interest had the same direction of effect with respect to the overall differential expression signatures.

### Machine learning framework

To systematically evaluate the clinical relevance of identified factors and validate their ability to encode information related to key outcomes, we developed supervised machine learning models. We trained models to predict three outcomes: presence of shock (defined as base deficit > 6), critical injury severity (ISS ≥ 25), and 28-day mortality. For each outcome, we developed parallel models using either gene expression data or derived factors as predictive features. For each input-outcome combination, we evaluated three classifiers: Lasso (LogisticRegression with L1 penalty; scikit-learn^32^), Random Forests (RandomForestClassifier; scikit-learn), and XGBoost (XGBClassifier; XGBoost^33^). For the Lasso models, we additionally scaled and centered the input features using scikit-learn’s StandardScaler.

We randomly partitioned the patients into training (80%) and held-out test (20%) sets using scikit-learn’s^32^ train_test_split function with stratified sampling to maintain outcome class distributions. Within the training set, we optimised model parameters using cross-validation. Specifically, we performed stratified 4-fold cross-validation (StratifiedKFold) and performed hyperparameter optimisation with Optuna^34^. We used RandomSampler for 100 trials, followed by 100 trials of TPESampler. We selected the models that maximise the mean cross-validated area under the receiver operating characteristic curve (AUC-ROC). Class imbalance was addressed by using balanced class weights for the Lasso and Random Forest models and by setting the scale_pos_weight argument of the XGBoost classifiers to the ratio of negative to positive samples. Additionally, we determined optimal classification thresholds within the validation folds using Youden’s J statistic, and applied this to calculate F1 scores.

Final model performance was assessed on the held-out test set using predictions averaged across the four cross-validation models. We also benchmarked these models against scikit-learn’s DummyClassifier using both the “stratified” and “most_frequent” strategies as baseline classifiers. To quantify feature importance and interpret model predictions, we employed SHAP (SHapley Additive exPlanations)^35^ analysis. For tree-based models, we used the TreeExplainer implementation, averaging SHAP values across the four cross-validation models to ensure robust feature ranking. This analysis provided a systematic framework for ranking genes and factors by their contribution to outcome prediction.

### Assessing module preservation

We used Weighted Gene Co-expression Network Analysis (WGCNA)^36^ to identify gene modules in the human and pig datasets. For the human dataset, a gene-gene correlation matrix was constructed using the biweight midcorrelation measure. To ensure that the network approximated scale-free topology, the function pickSoftThreshold was applied and a soft thresholding power of 10 was selected at which the scale free topology model fit (signed R²) reached 0.85. An adjacency matrix was computed from the similarity matrix using a signed network approach, and the corresponding Topological Overlap Matrix (TOM) was calculated. Hierarchical clustering based on the TOM was then performed, and modules were identified using dynamic tree cutting^20^ with a deepSplit parameter of 4, a minimum module size of 20, and a cut height of 0.975. Modules were subsequently merged if their eigengenes were highly correlated (mergeCutHeight = 0.07). Module were then refined by calculating module membership (kME) values. Genes initially labelled as “grey” (i.e. unassigned) were re-examined, those with a maximum kME value greater than 0.30 were reassigned to the module which they had the highest correlation. Genes whose intramodular kME was more than 0.10 lower than their maximum kME in another module were also reassigned. For the pig dataset, a custom function was used to compute a correlation matrix (rmcorr^37^), that is appropriate for repeated measures data^38^. Subsequent network construction and module refinement steps were similar to those applied to the human dataset.

To assess module preservation between species, analyses were restricted to the common genes present in both datasets. Pig Ensembl gene IDs were translated to their human orthologs using the getLDS function in biomaRt^10^. When multiple human mappings were observed for a single pig gene, the best match was selected based on orthology confidence and percent identity. Using WGCNA’s modulePreservation function^39^ with 400 permutations, composite preservation statistics (medianRank and Zsummary) were computed. Thresholds of Z = 2 and Z = 10 were applied to distinguish between low, moderate, and strong preservation, modules with Zsummary > 10 were considered strongly preserved.

To visualize the gene co-expression networks, we focused on the top hub genes by selecting the top 30 based on kME values and computing their correlation matrix. An absolute correlation threshold (> 0.75) was applied to form an adjacency matrix, which was then used to build a weighted, undirected graph using igraph^40^, which was subsequently visualised using ggraph.

### Pathway enrichment analysis

We performed enrichment analyses using the Molecular Signatures Database (MSigDB) C2 canonical pathways^41^, excluding the Chemical and Genetic Perturbations (CGP) subcategory. All pathway enrichment analyses were conducted using the clusterProfiler^42^ package.

For ranked gene lists, including differential expression results, treatment signatures and factors, we employed GSEA. We set minimum gene set size to 10 and performed 10,000 permutations, with an epsilon value of 1×10^−50^. In the results section, we present a subset of the leading edge genes (LEGs) alongside pathways. For discrete gene sets, including shock-associated genes (n=396), temporal gene clusters, and weighted gene co-expression network analysis (WGCNA) modules, we instead performed overrepresentation analysis using the enrichr function^42^. For overrepresentation analyses, we excluded pathways with fewer than three genes overlapping with the gene set.

All p-values were adjusted for multiple testing using the Benjamini-Hochberg procedure, with significance determined at 5% false discovery rate (FDR). To address redundancy among the 2,982 tested pathways, we employed the SimplifyEnrichment package^43^ to cluster similar pathways based on shared genes. For GSEA results, clustering was performed using LEGs, while for overrepresentation analyses, we used genes present in both the pathway and input gene set. Pathway clusters were defined using the binary cut algorithm with default parameters. For visualisation, we present the most statistically significant pathway from each cluster as a representative example.

## Supplementary Figures

**Supplementary Figure 1.**
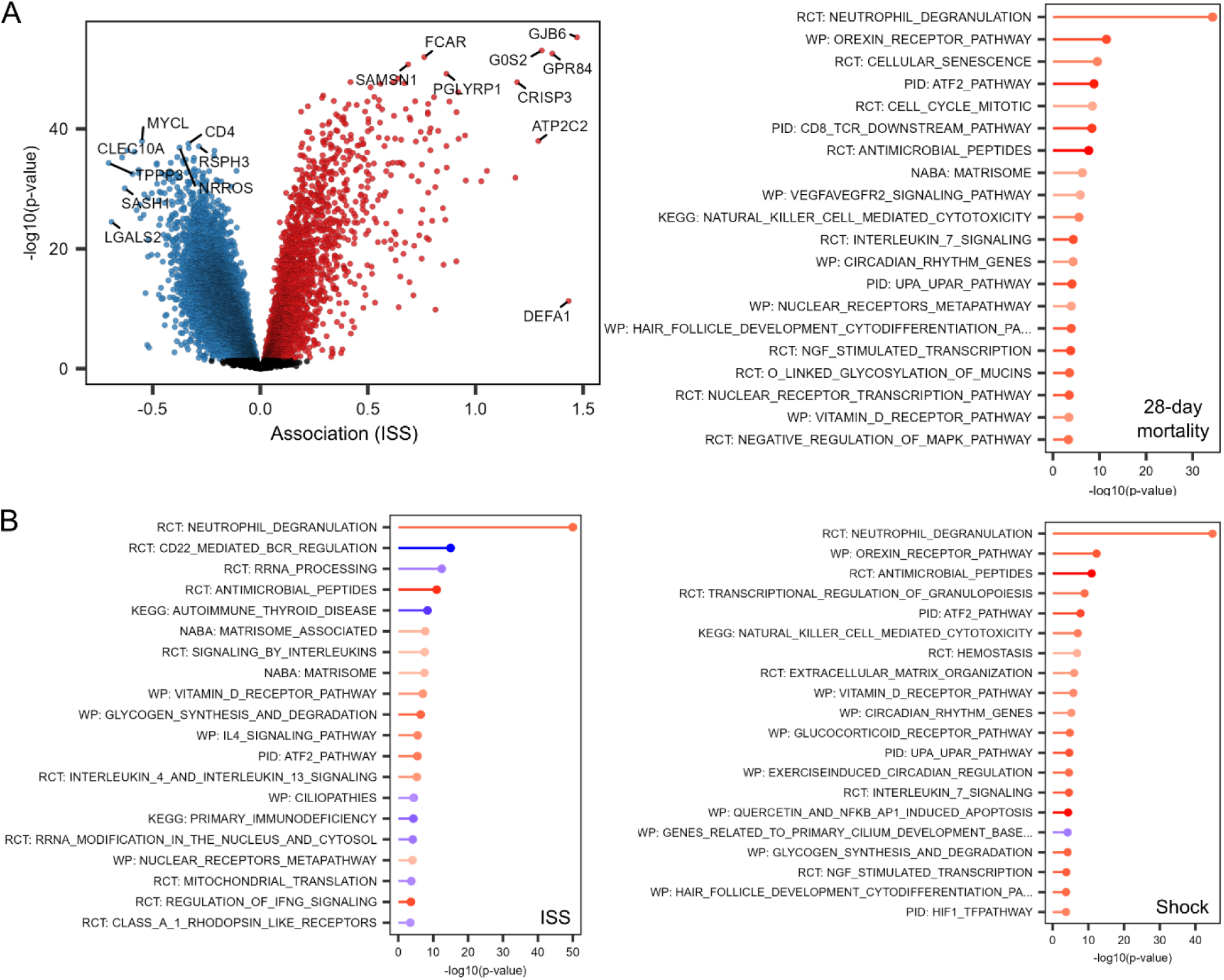
Genes and pathways associated with outcomes for human patients. **A**) Volcano plots displaying gene associations with ISS. Upregulation indicates higher gene expression for patients with greater ISS scores. **B**) The 20 most significantly enriched pathways for each of the three human outcomes (ISS, shock and survival), ordered by p-value and coloured by enrichment score. Positive scores (red) indicate upregulation, and negative scores (blue) indicate downregulation.

**Supplementary Figure 2.**
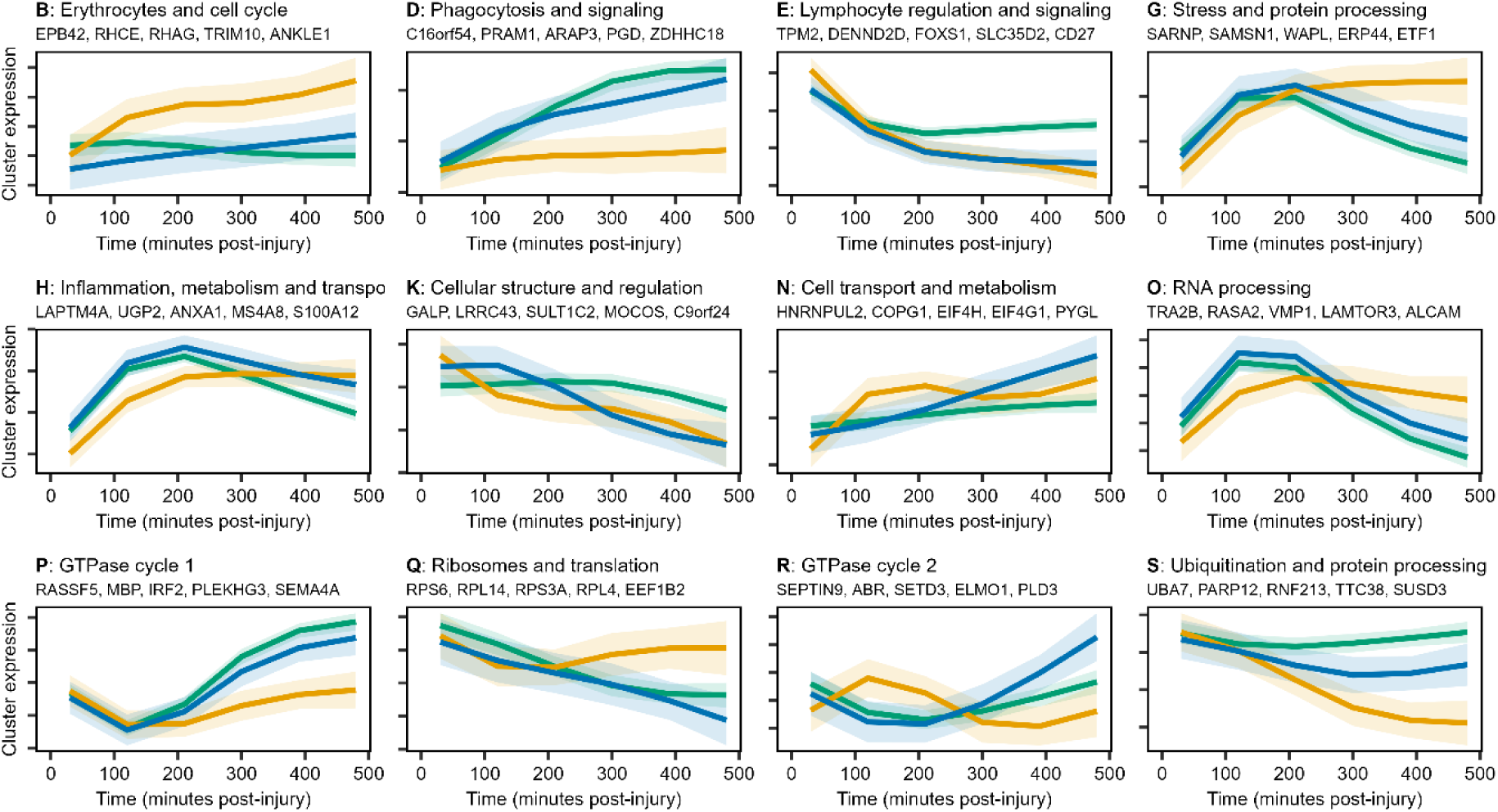
Representative expression patterns for clusters of genes with similar temporal profiles. Genes are shown that were not included in panel C of **Figure 4**. Lines, shaded areas and colour indicate estimated means, 95% confidence intervals and treatment group, respectively. Plot titles indicate the cluster letter, name and five key genes.

**Supplementary Figure 3.**
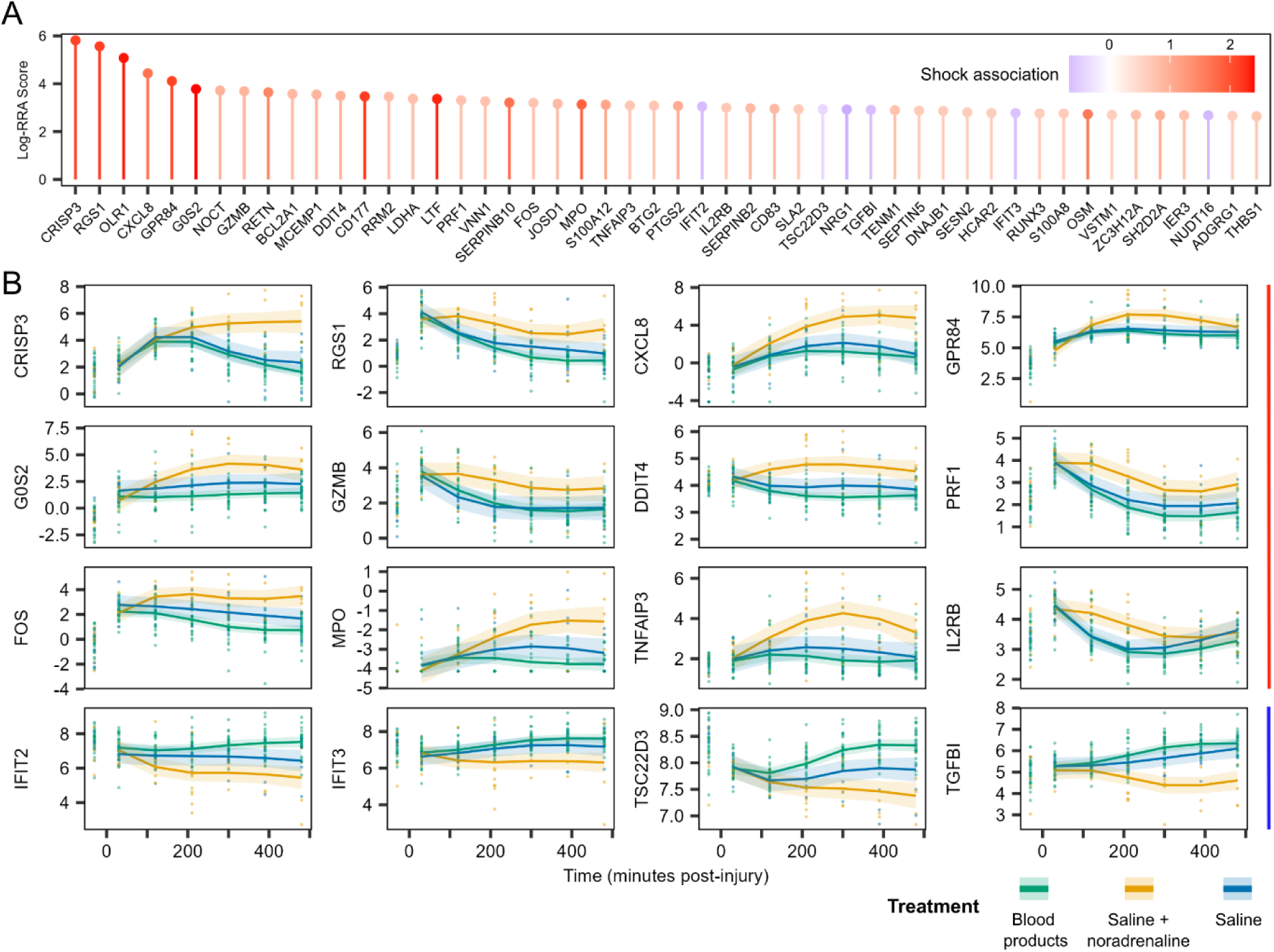
Genes associated with shock and modified by treatment. **A**) The 50 genes with the highest log-RRA scores, where greater scores indicate stronger associations with shock (humans) and greater differences between treatment groups across time (porcine model). Colour indicates association with shock (log-fold change) in human patients, where positive values (red) indicate positive associations, and negative values (blue) negative associations. **B**) The temporal profiles of example genes over time in the porcine model, stratified by treatment group. Lines and shaded areas represent estimated means and 95% confidence intervals, respectively. Points indicate underlying data and colours indicate treatment group.

**Supplementary Figure 4.**
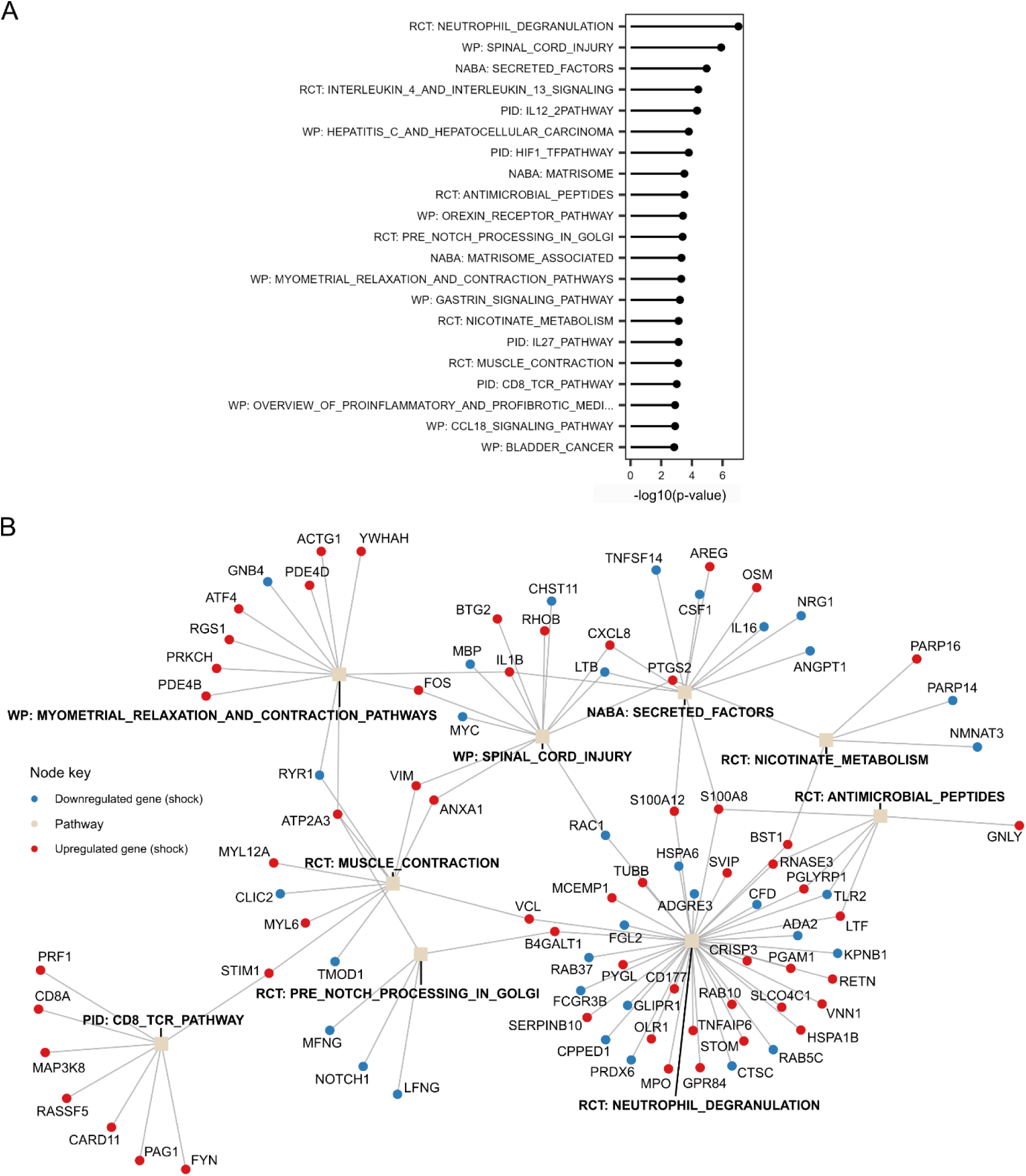
Pathways associated with the genes associated with shock and treatment. **A**) The 20 pathways most significantly enriched for the 396 genes associated with shock in humans and treatment in the porcine model. −log10-transformed p-values are provided based on the results of a hypergeometric test. **B**) Network connecting pathways and the genes that belong to them. The top 10 most significantly enriched pathways are shown for the set of 396 genes that were significantly associated shock in humans and treatment in the porcine model. The member genes are displayed for each pathway, with colour indicating whether these genes are up- or down-regulated in shock.

**Supplementary Figure 5.**
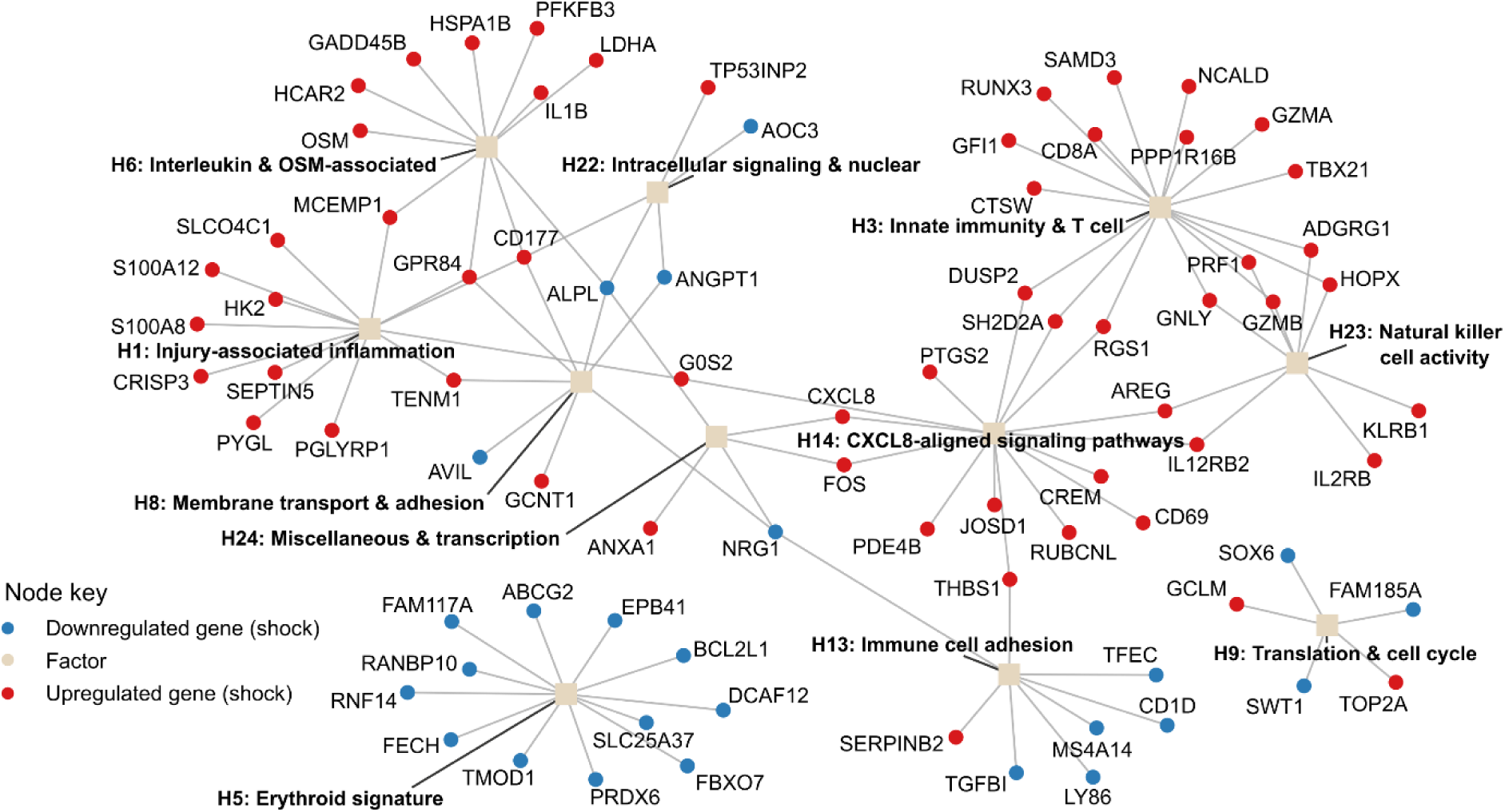
Network connecting genes associated with shock and treatment type to factors. Factors are shown that were connected to at least 5 of the 396 genes that were significantly associated with shock in humans and treatment in the porcine model.

**Supplementary Figure 6.**
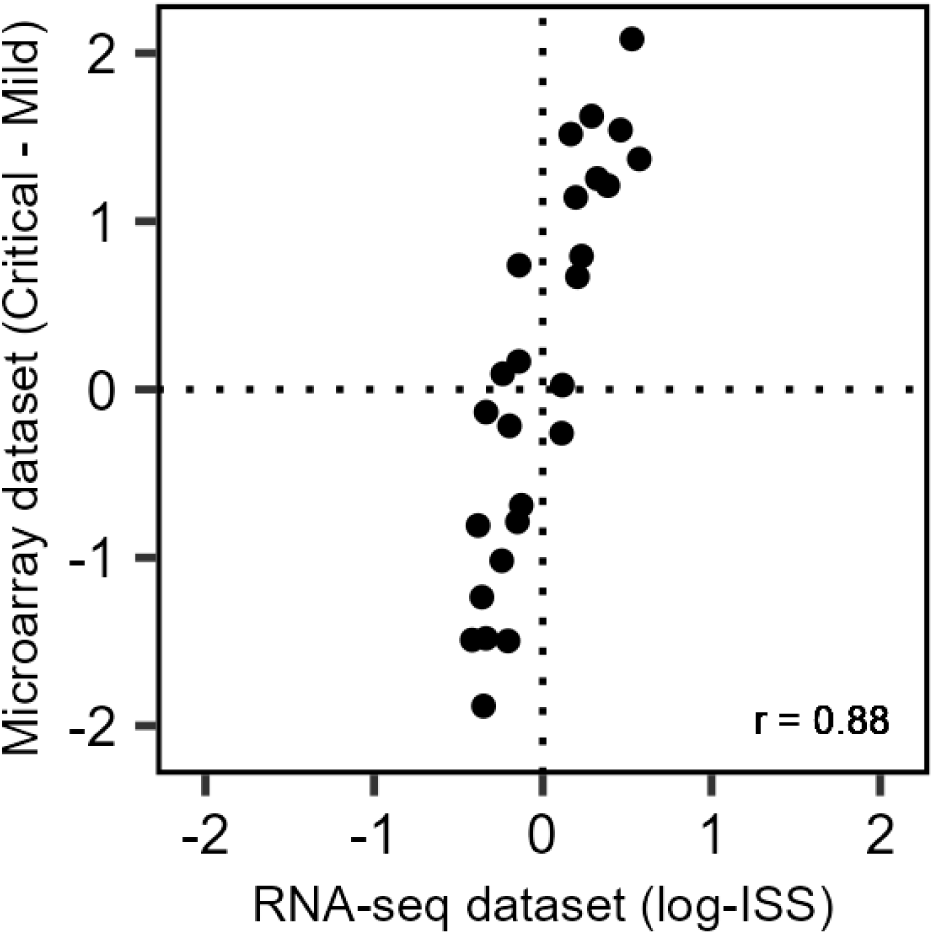
Comparing the effect of injury on factors across datasets. The model estimates for the comparison of critical and mild patients in the external microarray dataset (y-axis) are compared to those for ISS in the human trauma dataset in a scatter plot. Dotted lines indicate 0 estimates for reference. Correlation (Pearson’s r) is shown for the data points.

**Supplementary Figure 7.**
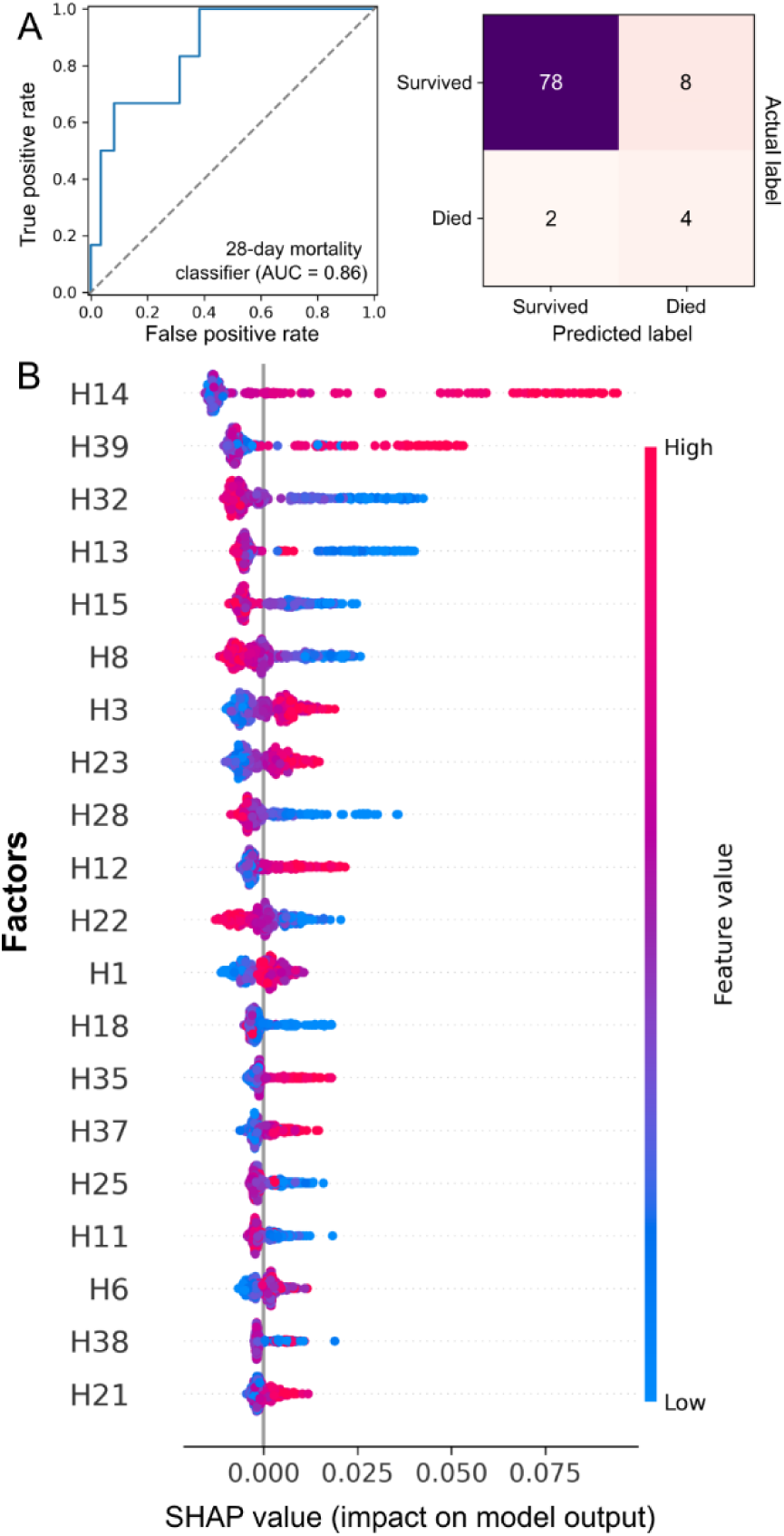
**A**) Receiver operating characteristic (ROC) curve (left) and confusion matrix (right) for an XGBoost model developed to predict 28-day mortality. The area under the curve is displayed. **B**) SHapley Additive exPlanation plots indicating the impact of factors on the prediction of 28-day mortality. Colour indicates the feature value (i.e., high or low factor expression). SHAP values indicate impact on the output, with higher values increasing the likelihood of predicting a patient as dying within 28-days of injury (right).

**Supplementary Figure 8.**
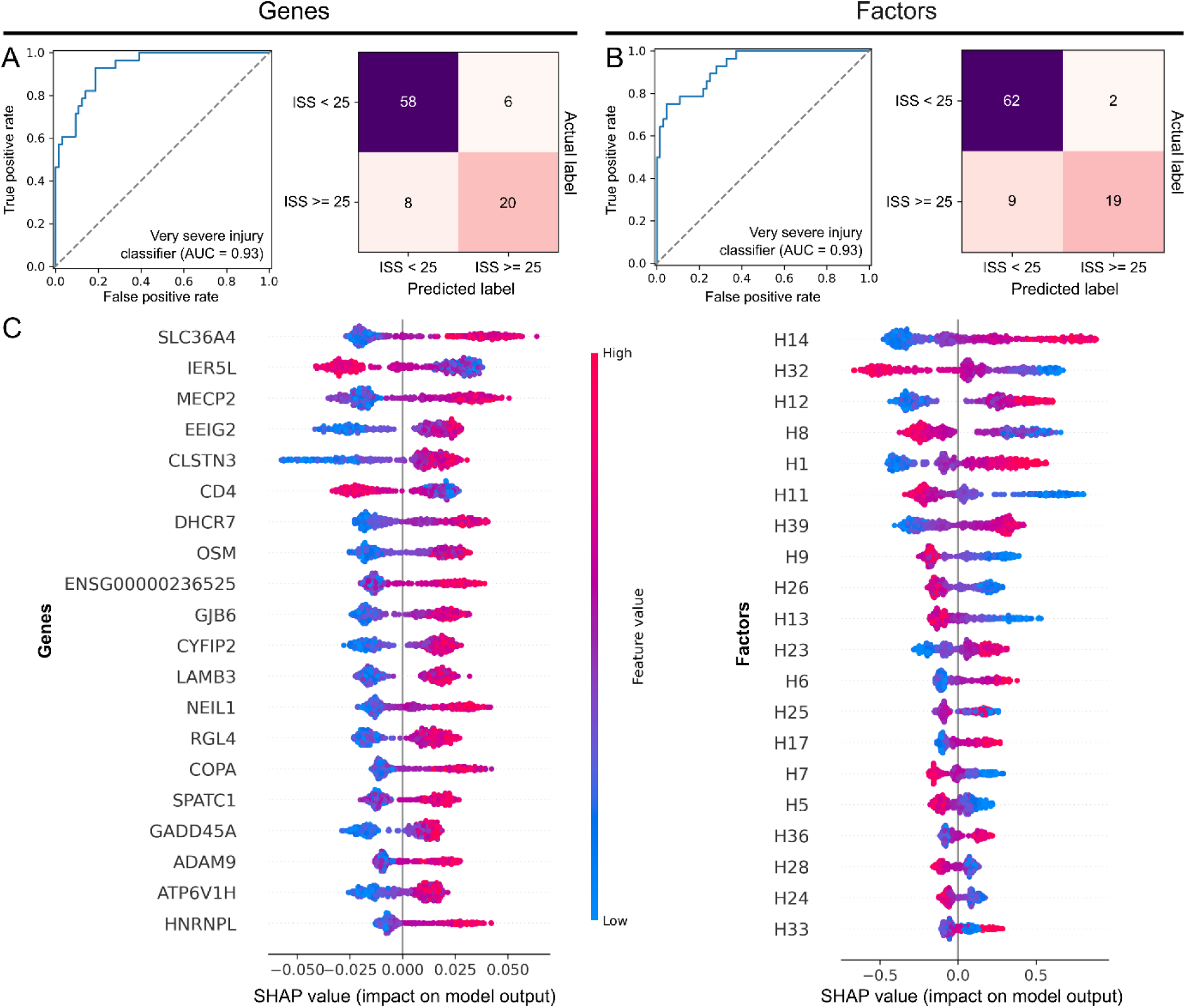
Predicting severe injury using XGBoost. Model performance summaries and SHAP plots for models trained to predict whether a given sample had an ISS score greater than 24. **A/B**) Receiver operating characteristic (ROC) curves (left) and confusion matrices (right) are shown for each model. The area under the curve is displayed. Plots display predictive performance for the factor-based model (**A**) and the model trained on the original gene expression data (**B**). **C**) SHAP plots indicating the impact of genes (left) and factors (right) on the prediction of critical injury. Colour indicates the feature value (i.e., high or low gene expression). SHAP values indicate impact on the output, with higher values increasing the likelihood of predicting a patient as having severe injury.

**Supplementary Figure 9.**
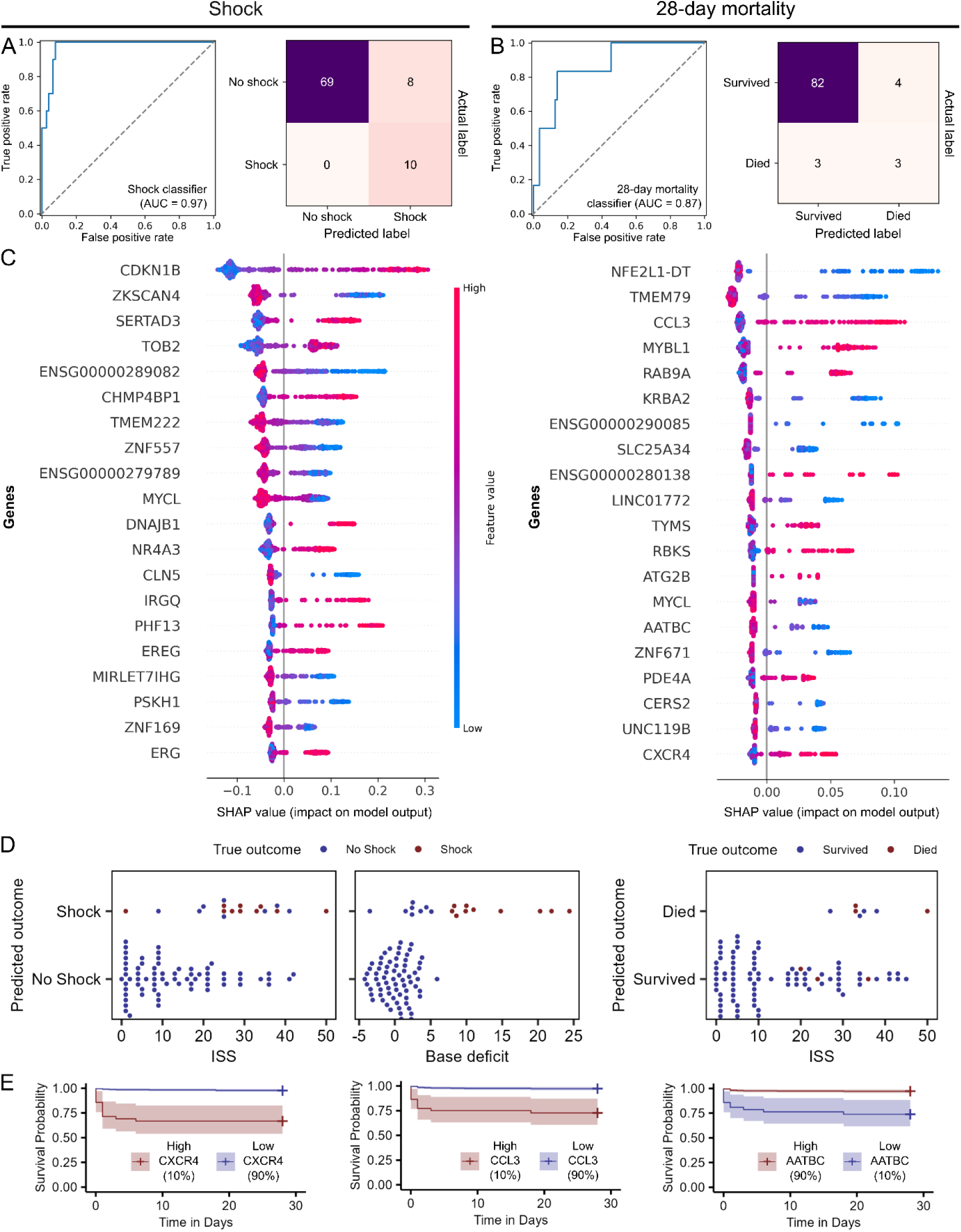
Predicting shock and survival from gene expression using XGBoost. **A/B**) Receiver operating characteristic (ROC) curves (left) and confusion matrices (right) are shown for each model. The area under the curve (AUC) is displayed. Plots display predictive performance for the shock model (**A**) and the 28-day mortality model (**B**). **C**) SHapley Additive explanation (SHAP) plots indicating the impact of genes on the prediction of shock (left) and 28-day mortality (right). Colour indicates the feature value (i.e., high or low gene expression). SHAP values indicate impact on the output, with higher values increasing the likelihood of predicting a patient as having shock (left) or dying within 28-days of injury (right). **D**) Beeswarm plots displaying ISS score and base deficit as a function of the true and predicted class. Distributions shown for the prediction of shock (left) and 28-day mortality (right). **E**) Survival curves stratified based on the expression of genes in the human cohort.

**Supplementary Figure 10.**
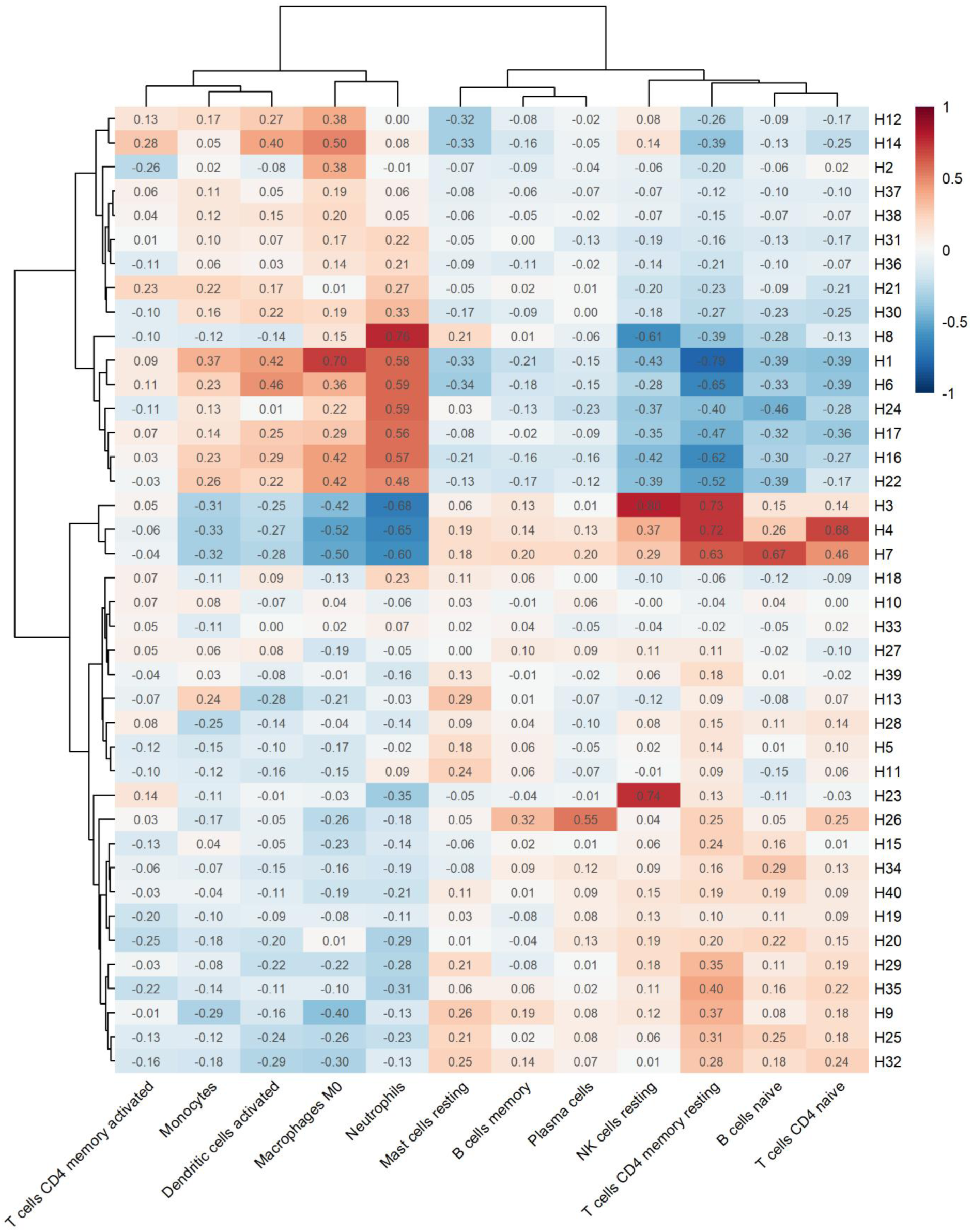
Correlations of factors with imputed immune cell abundances. Heatmap displaying Pearson’s r (numbers and colour) comparing the expression of factors to the abundances of immune cells predicted using CIBERSORTx. Cell types with an estimated abundance of 0 for more than 50% of samples were removed. Rows and columns are ordered by hierarchical clustering.

**Supplementary Figure 11.**
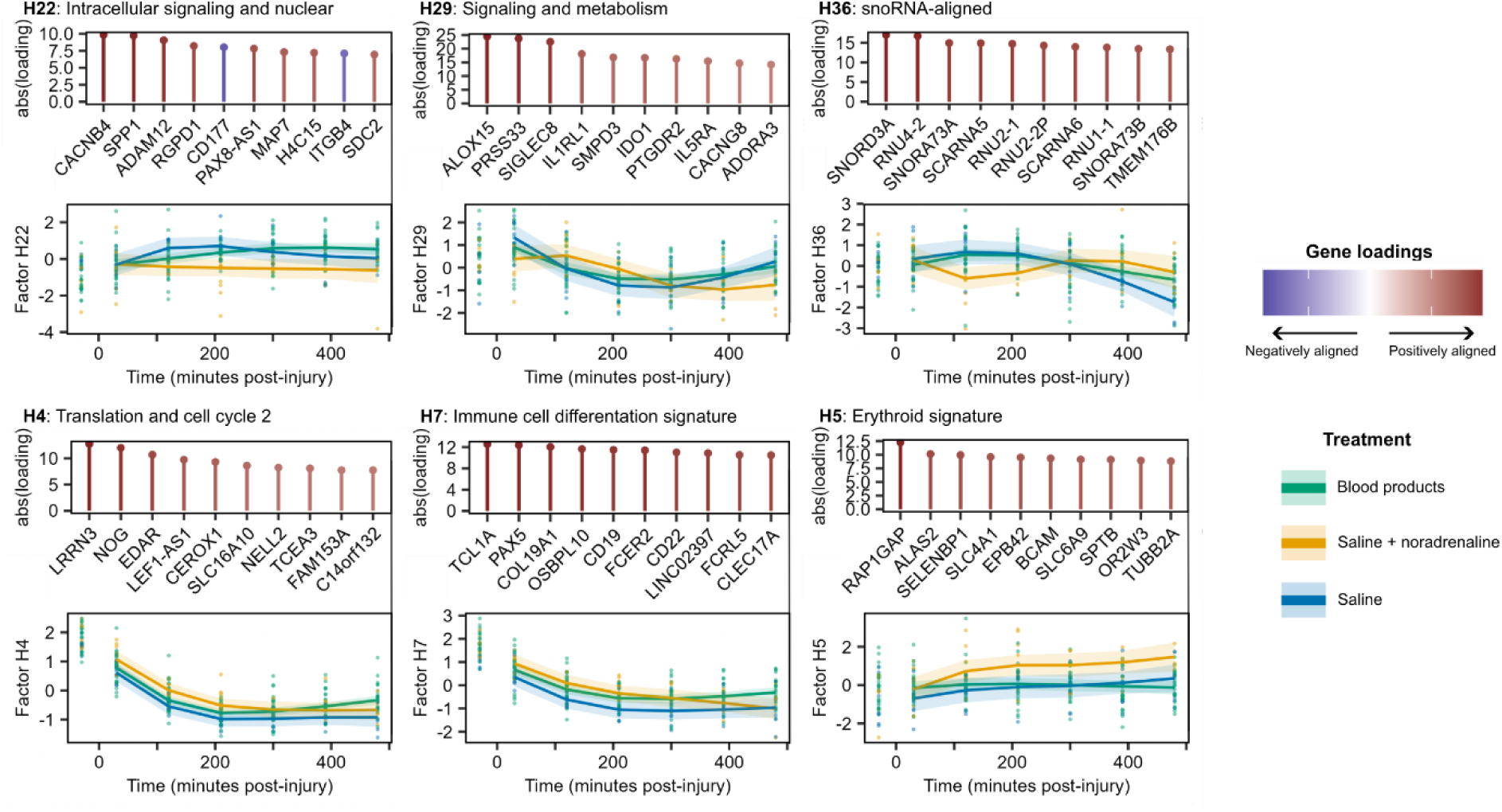
Summaries of other factors with TxT profiles. Factors are shown that had i) significant TxT interactions (5% FDR), ii) were associated with a human outcome (5% FDR), iii) were associated with at least one pathway (5% FDR) and iv) were not shown in **Figure 6**. Summaries include the factor ID and name (**Table S17**) of the factor in the porcine model. The temporal expression plots show estimated means (lines) and 95% confidence intervals (shaded areas) of factor expression in the porcine data, stratified by treatment group, with points indicating underlying data.

**Supplementary Figure 12.**
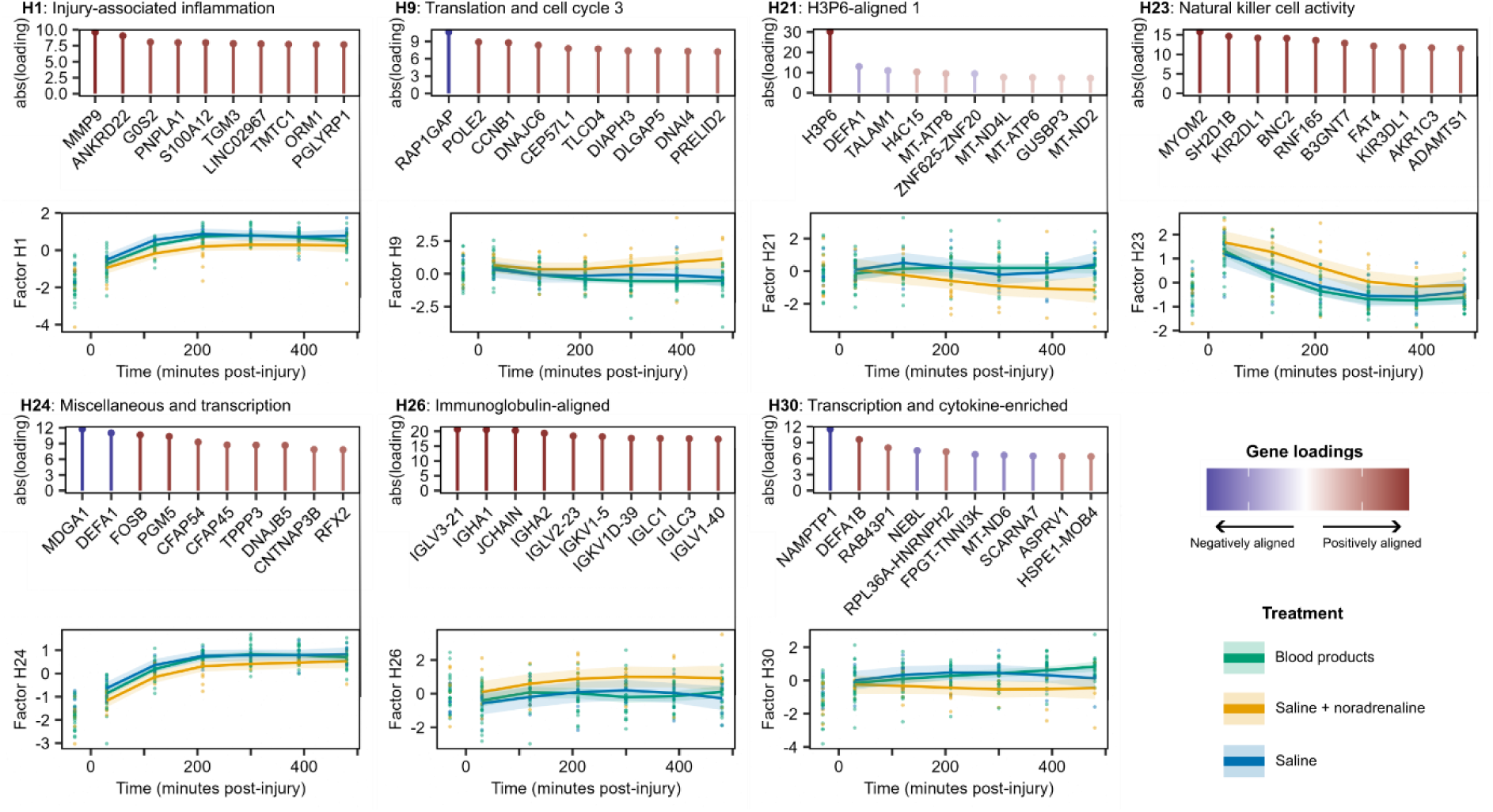
Summaries of factors with consistent effects of treatment. Factors are shown that i) had significant main effects of treatment (5% FDR), ii) were associated with a human outcome (5% FDR) and iii) were associated with at least one pathway (5% FDR). Summaries include the factor ID and name (**Table S17**), the top three enriched pathways, the loadings for the most highly aligned genes and the estimated expression of the factor in the porcine model. The temporal expression plots show estimated means (lines) and 95% confidence intervals (shaded areas) of factor expression in the porcine data, stratified by treatment group, with points indicating underlying data.

**Supplementary Figure 13.**
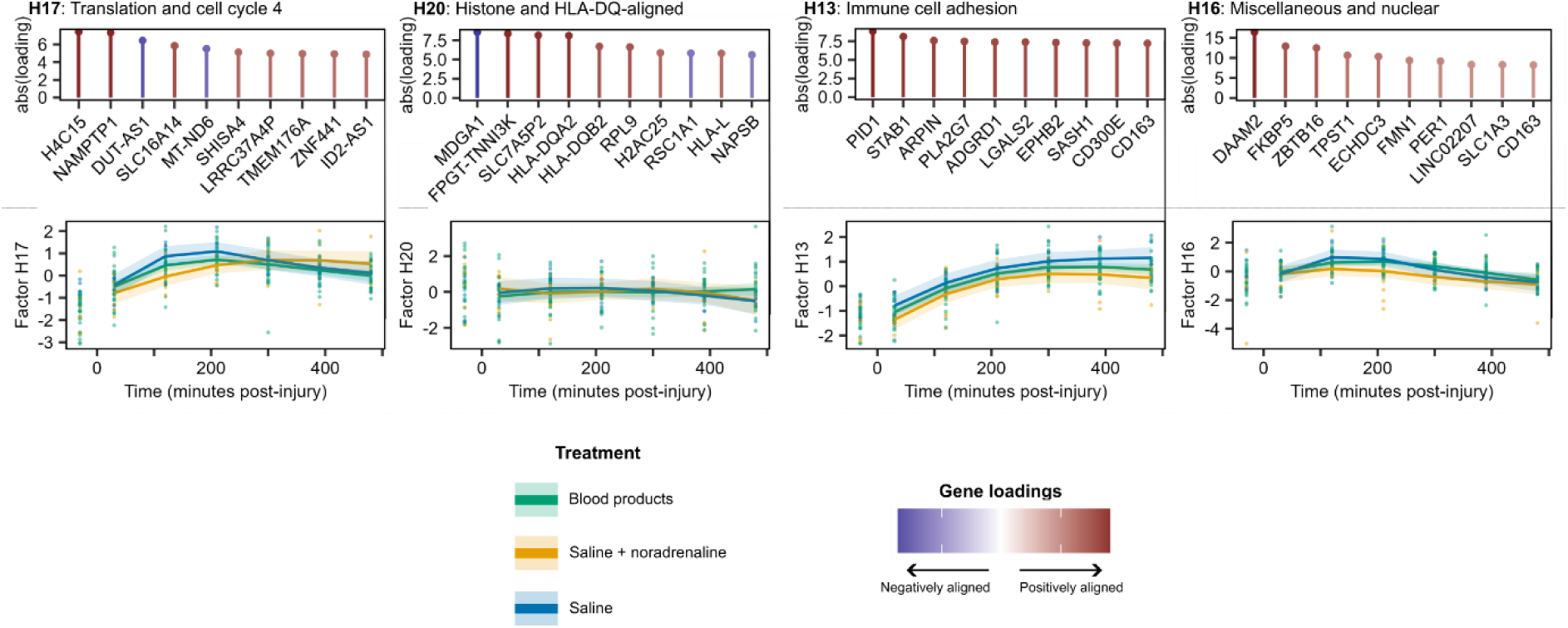
Summaries of other factors with no significant treatment differences. Factors are shown that were i) not associated with treatment type (5% FDR), ii) associated with a human outcome (5% FDR), iii) associated with at least one pathway (5% FDR) and iv) not shown in **Figure 6**. Summaries include the factor ID and name (**Table S17**), the top three enriched pathways, the loadings for the most highly aligned genes and the estimated expression of the factor in the porcine model. The temporal expression plots show estimated means (lines) and 95% confidence intervals (shaded areas) of factor expression in the porcine data, stratified by treatment group, with points indicating underlying data.

**Supplementary Figure 14.**
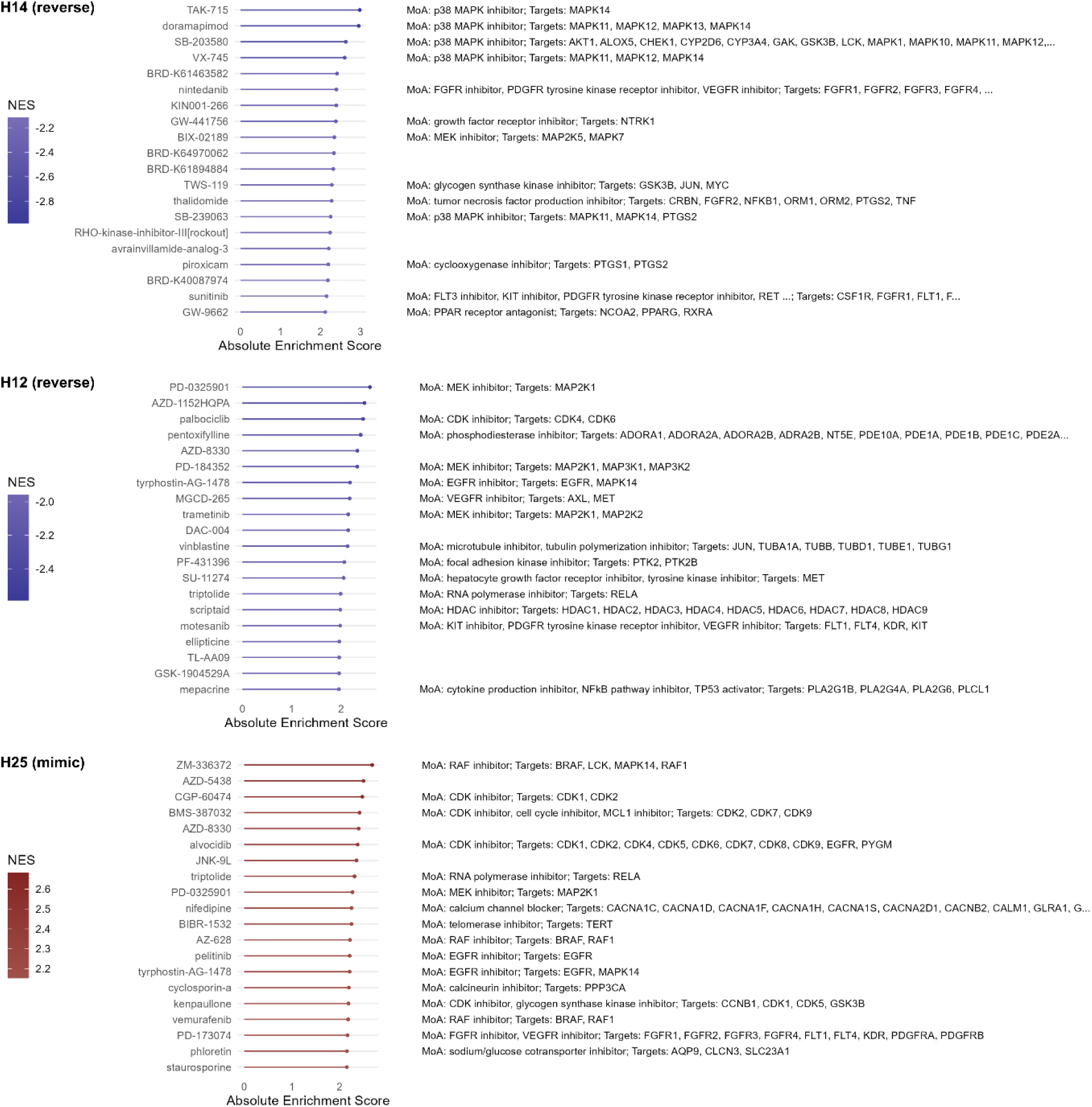
Alignment of factors with drug perturbations. Dotplots indicate the enrichment of drug perturbations for factors H14, H12 and H25. Negatively aligned (i.e., reverse) perturbations are shown for H14 and H12, since they were upregulated in shock. Conversely, H25 was downregulated in shock, so positively aligned (i.e., mimic) perturbations are displayed for H25. The top twenty significantly enriched (5% FDR) perturbations are shown, based on NES score. The MoA(s) and target(s) are shown alongside each perturbation, where available.

**Supplementary Figure 15.**
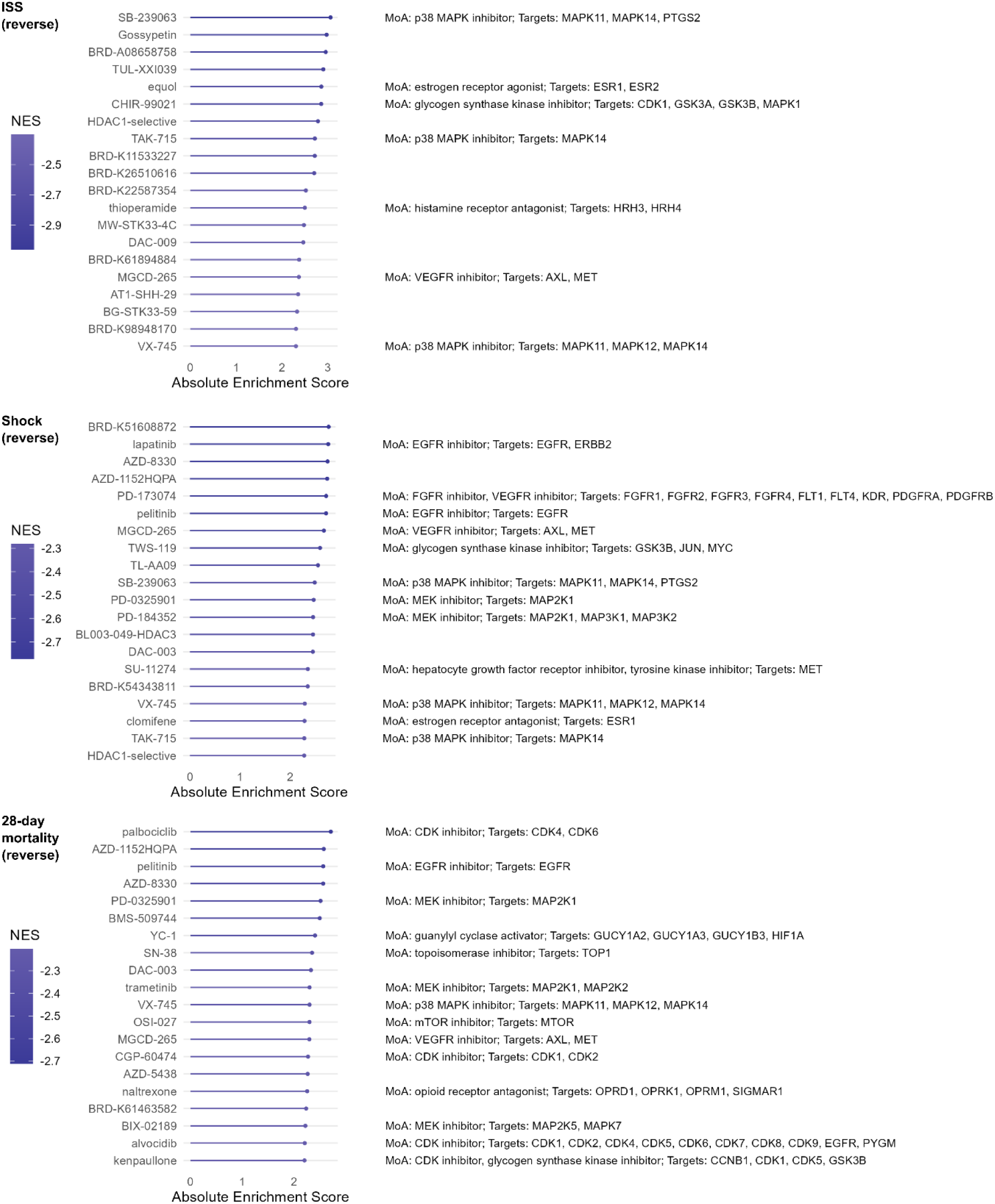
Alignment of human outcomes with drug perturbations. Dotplots indicate the enrichment of drug perturbations for each human outcome. Only negatively aligned (i.e., reverse) perturbations are shown. The top twenty significantly enriched (5% FDR) perturbations are shown, based on NES score. The MoA(s) and target(s) are shown alongside each perturbation, where available.

**Supplementary Figure 16.**
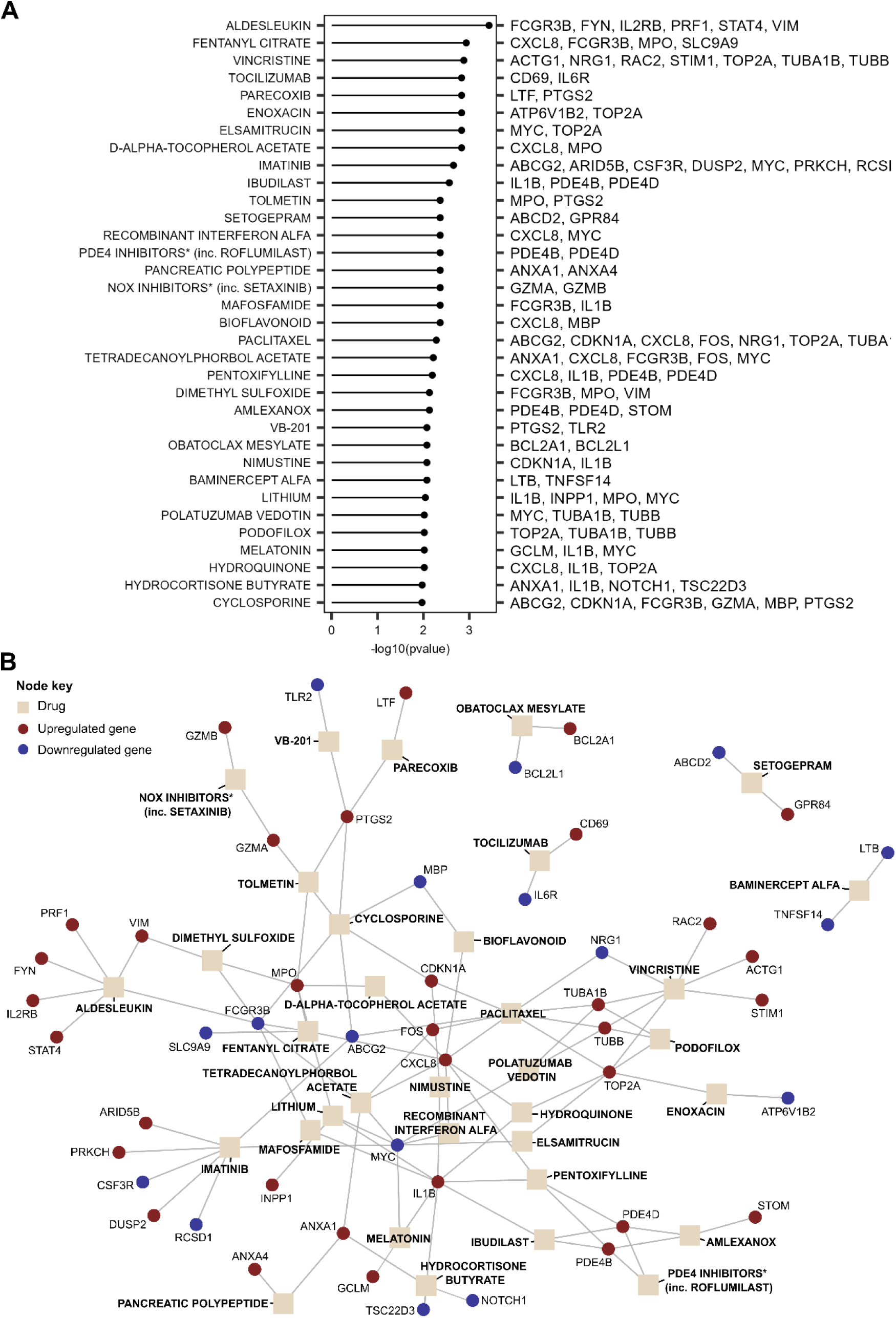
Drugs interacting with shock- and treatment-associated genes. Drugs are shown whose targets were overrepresented in the 396 genes associated with shock in humans and treatment in porcine models, according to a fisher’s exact test (5% FDR). **A**) Barplot showing, for each drug, the p-value (left) and the genes that interact with that drug (right). *In some cases, there were a number of drugs linked to the same set of genes; in these cases, we report the drug class, with an asterisk and the first drug in the category. **B**) Network plot displaying interactions (edges) between drugs (beige squares) and genes (green circles) recorded in DGIdb.

